# Resonance sonomanometry for noninvasive, continuous monitoring of blood pressure

**DOI:** 10.1101/2023.12.24.23300502

**Authors:** Raymond Jimenez, Dominic Yurk, Steven Dell, Austin C. Rutledge, Matt K. Fu, William P. Dempsey, Yaser Abu-Mostafa, Aditya Rajagopal, Alaina Brinley Rajagopal

## Abstract

Cardiovascular disease is the leading cause of death worldwide. Existing methods for continuous, noninvasive blood pressure monitoring suffer from poor accuracy, uncomfortable form factors, or a need for frequent calibration, limiting their adoption. We introduce a new framework for continuous BP measurement that is noninvasive and calibration-free. The method uses acoustic stimulation to induce resonance of the artery wall. Ultrasound imaging is used to measure resonance and capture arterial dimensions which are related to blood pressure via a fully-determined physical model. The approach and model are validated *in vitro* using arterial mock-ups and then in multiple arteries in human subjects. Further development could facilitate more robust continuous blood pressure measurement, providing significant benefits for early diagnosis and treatment of cardiovascular disease.

## Introduction

Blood pressure (BP) is a critical metric for clinicians when assessing patient health, and as such, continuous, noninvasive measurement has been of long-term interest to the scientific and clinical communities. In an artificial system, fluid pressure would normally be assessed by simply placing a pressure transducer inside the vessel of interest. A similar approach can be applied to human arteries by inserting an arterial catheter, which produces continuous and accurate BP measurements. These continuous waveforms provide significant clinical value which cannot be obtained solely from maximum (systolic) and minimum (diastolic) BP values, including diagnosis of conditions such as aortic stenosis and valve insufficiency (*1*), measurement of cardiac efficiency (*2*), and early identification of hypotensive crises (*3*). However, arterial catheterization is an invasive procedure which is time consuming and carries risks of pain, infection, hemorrhage, and ischemia (*4*). As a result, it is typically only performed in critical care units and operating rooms. In all other situations, BP is measured using an inflatable arm cuff, which operates by applying external pressure until the artery collapses (i.e., blood flow stops), and the measurement is taken when pressure is gradually released and blood flow resumes. While these cuffs are noninvasive, they only provide intermittent and often inaccurate measurements compared to measurements from invasive catheters (*5–7*), and repeated cuff inflation can cause significant discomfort for patients (*8*).

The shortcomings of inflatable BP cuffs have inspired the development of numerous continuous, noninvasive BP (cNIBP) measurement techniques based on a variety of physical signals and phenomena (*9*). These include measurement of fingertip blood perfusion (volume clamping) (*10*), pressure signals at the surface of the skin (tonometry) (*11, 12*), light reflectance from blood (photoplethysmography) (*13*), electrical conductivity of blood (bioimpedance) (*14*), pressure wave velocity (pulse transit time) (*15*), reactive forces from cardiac ejection (ballistocardiography) (*16*), and blood velocity (Doppler ultrasound) (*17–21*). However, most of these methods require calibration against an inflatable cuff which must be repeated regularly due to dynamic change in BP and arterial physiology (*22, 23*). Methods that do not require calibration carry drawbacks such as periodic data blackouts (*24*) or reliance on “black-box” machine learning techniques that may overfit the demographics of the underlying dataset (Table S1 and Supplementary Text 2.1) (*25*). These constraints significantly limit the ability of these methods to report accurate blood pressure in dynamic conditions such as those found in critically ill patients, where vital signs can rapidly change.

Here, we present a new method for cNIBP measurement based on the phenomenon of resonance sonomanometry (RSM) (Fig. 1, Fig. S1). With this method, the artery is stimulated by an acoustic transducer while its resonant response and dimensions are simultaneously measured using ultrasound imaging (Fig. 1A, B). Our motivation comes from the tuning of a guitar string: change the tension in a string, and its resonant tone changes proportionally to tension applied. By plucking the string and measuring the resonant frequency, this relationship can be inverted to calculate the absolute tension applied. We extend this analysis to the circumference of the artery (Fig. 1C). Circumferential wall tension is directly related to fluid pressure inside the artery per Laplace’s law, and the specific value of tension can be inferred from the artery’s dimensions and resonant frequency. This pressure-resonance relationship is the unique, defining characteristic of the method: applying acoustic stimulus allows determination of the artery’s resonant frequency, exposing sufficient information for determination of absolute pressure in an arterial system. This frequency information removes the need for any calibration or external reference.

**Figure 1:**
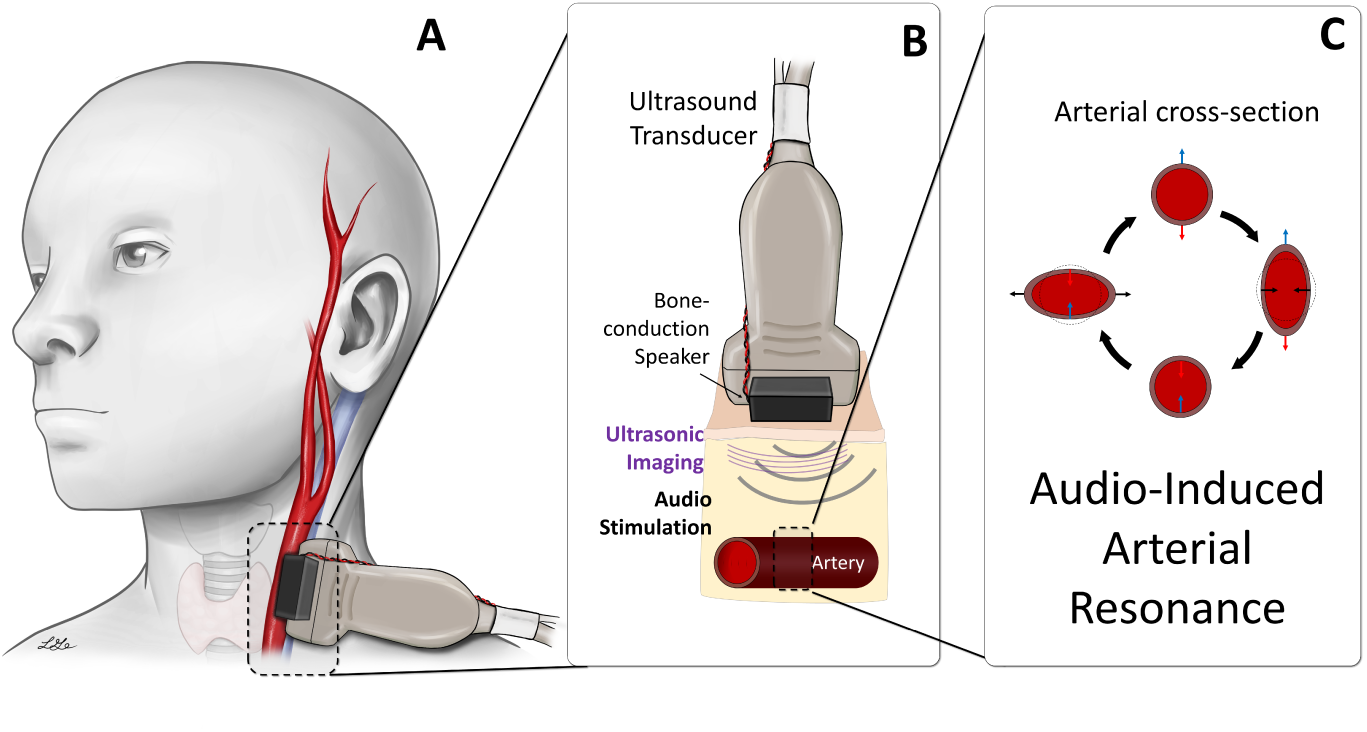
Acoustic stimulation paired with ultrasound imaging reveals resonance properties of an artery. (**A**) Device placement to measure blood pressure in the carotid artery. (**B**) Illustration of device operation: Ultrasound (gray probe) is used to generate images of the artery (bottom). The device provides acoustic stimulus via moving-coil drivers (black boxes) which induce arterial response. (**C**) Illustration of vascular resonance response (VRR). If the frequency of the stimulus aligns with the resonant frequency of the artery, the artery’s cross-section oscillates between two forms (middle-left and middle-right), exaggerated for clarity. The top and bottom walls of the artery move relative to the ultrasound probe (motion toward and away from the probe are indicated as blue and red, respectively), which is captured by ultrasound imaging. Differential velocity of the walls (top velocity minus bottom velocity) is calculated. Comparing differential wall velocities over a range of stimulus frequencies enables identification of the resonant frequency.

This model is validated through measurements of an arterial mock-up over a physiologically relevant range of pressures and dimensions. We then show through initial human tests that resonance can be stimulated and can be used to generate BP waveforms on central (carotid) and peripheral (axillary, brachial, and femoral) arteries. These measurements show that the resonance predicted by the physical model not only persists in much more complex *in vivo* systems, but also that the method produces data with high time resolution, providing continuous, calibration-free measurements of the full absolute (not relative) BP waveform.

### Background on the physical model

The physical model draws from two disparate lines of analysis: one from aerospace engineering and the other from biomechanics. The first set of analyses deals with the acoustic resonance modes in thin-walled cylindrical shells for large-scale industrial applications such as fuel tanks and pipelines (*26*). The second set of analyses examines the dynamics of *in vivo* arterial walls by modeling them as long, thin-walled cylindrical shells and using structural and fluid mechanics to calculate how these shells respond to changes in pressure (*27, 28*). While these two lines of analysis share fundamental commonalities and assumptions, no work has combined them to create a model of the resonant modes in pressurized arteries. Furthermore, all of these analyses focus on deriving expected responses based on a known applied pressure. By combining and inverting these relationships, we demonstrate that it is possible to calculate *in vivo* arterial blood pressure from measurements of a vessel’s resonant response.

In order to extend the analysis of inanimate objects (*26*) to the *in vivo* context, we used additional mechanical analyses to account for various physical complexities inherent to living systems. These included the presence of a pressurized fluid inside the artery (*29*) and inertial damping due to fluid mass inside and outside the artery (*30, 31*). Furthermore, we integrated established biomechanical analyses to account for effects such as the significant distention of the artery as pressure changes and the nonlinear character of its elasticity (*27, 28*).

## Results

### Physical model and direct calculation of pressure

Arteries are modeled as an elastic, thin-walled tube; the interior is filled with a fluid (blood) under pressure (with or without flow), and this tube is fixed in a surrounding fluid. We analyze this system and find that the final relation for calculating blood pressure (*P* ) depends on seven parameters: *a* (artery radius to the center of the wall), *h* (artery wall thickness), *f* (artery resonant frequency in Hz), *E* (arterial wall Young’s modulus), *ν* (arterial wall Poisson ratio), *ρ_S_* (arterial wall density), and *ρ_L_* (density of fluid surrounding the artery). The first two parameters are commonly measured today using ultrasound imaging, while the last three are nominally consistent across individuals and are estimated from the IT’IS material database for human tissues (*32*), leaving *f* and *E* as the remaining parameters to be determined. *f* is calculated by fitting our measured frequency response, and *E* is calculated using the relation 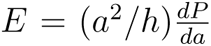, obtained from the Moens-Korteweg and Bramwell-Hill equations (*17*) (Fig. S2, Table S2, Supplementary Text 2.2). The full pressure equation is given in Equation 1 (see Materials and Methods for more on the physical model):

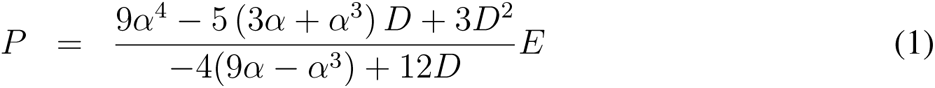

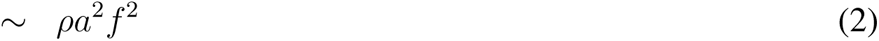

where

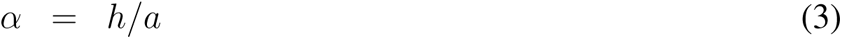

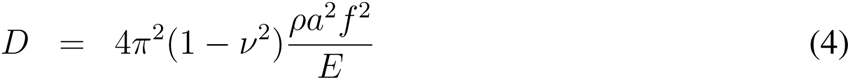

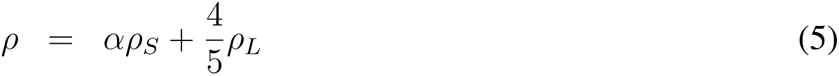

In these equations, *α* and *D* are dimensionless thickness and frequency parameters and *ρ* has units of mass density per unit volume. Equation 1 represents the inverted equation for pressure using the dominant frequency mode, while Equation 2 describes the system’s leading-order scaling. These relationships are applicable in the limit when the shell is thin-walled (i.e., *h ≪ a*, *α ≪* 1). In particular, the *P ∼ f* ^2^ scaling relation aligns with our intuition from the guitar string and is further substantiated by the sensitivity analysis (Supplementary Text 2.3); as blood pressure (and therefore wall tension) increases, so too does the resonant tone of the artery.

### Validation of physical model in arterial mock-ups

We first validated the pressure-resonance relationship *in vitro* using a custom device that combined ultrasound imaging and acoustic stimulation (Fig. 1B). We examined resonance in a long, cylindrical arterial mock-up (arterial imaging phantom) fabricated from thin-walled rubber tubing (Fig. 2A, B). First, we confirmed the presence of the resonance predicted by our model. We inflated the tubing with 75 mmHg of pressure, which is a clinically useful value and swept an acoustic stimulus across a wide range of frequencies while imaging the tubing with ultrasound (Fig. 2C). There were two prominent features in the measured frequency response that are characteristic of resonance: a large spike in magnitude space and a sigmoidal phase response with height *π* radians, both centered at the same frequency. The presence and sharpness of these features indicated that circumferential resonance was successfully stimulated and measured.

**Figure 2:**
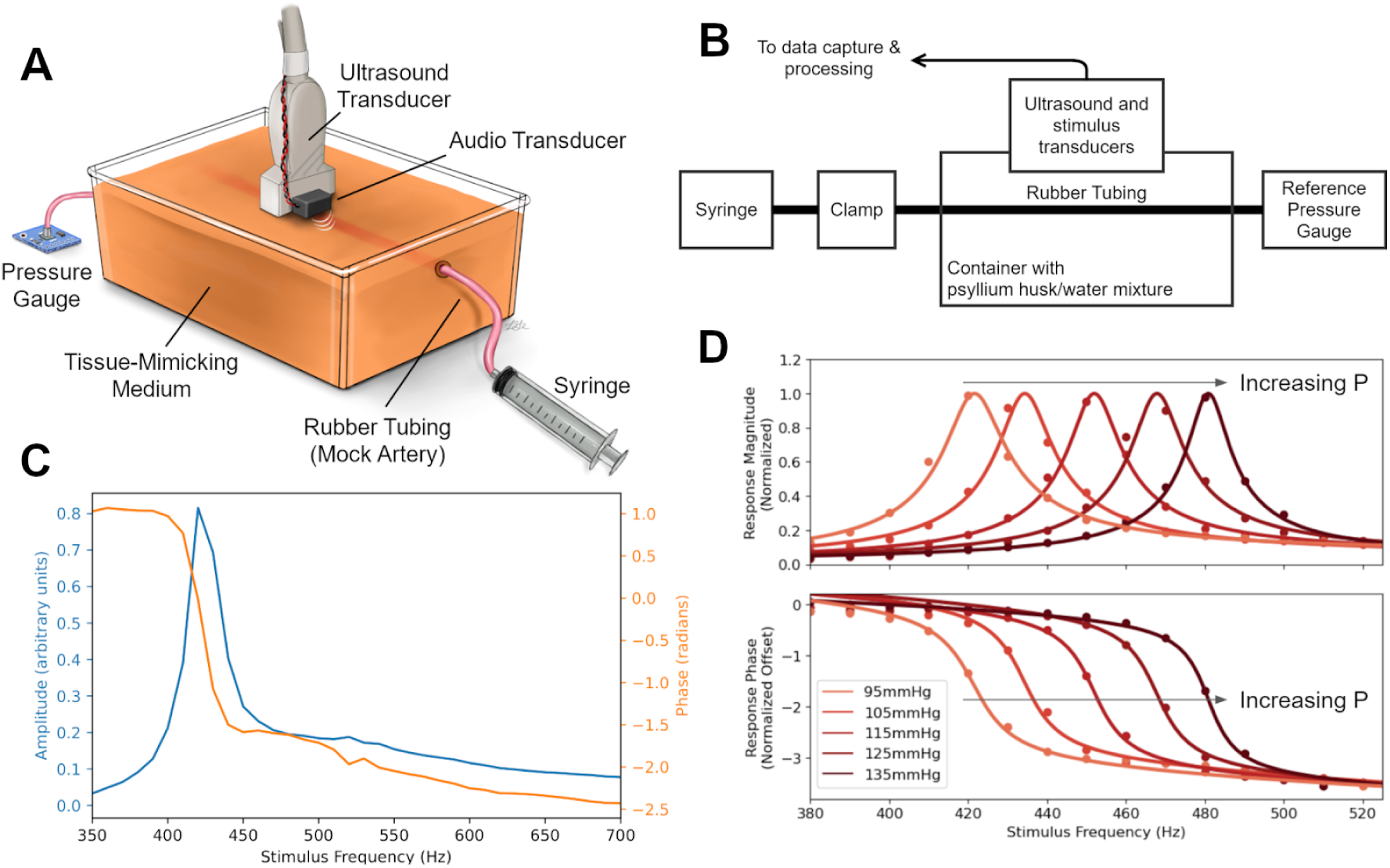
Resonance response in artery mock-up. (**A**) Illustration of mock artery, showing relative positioning of components. (**B**) System diagram, displaying connection between components. Thin latex rubber tubing filled with water connects a syringe and a digital pressure transducer. The syringe is depressed, increasing the pressure, and clamped once the desired pressure is reached. The rubber tubing is placed in a bath of psyllium husk/water mixture, which serves as a scattering medium to simulate surrounding tissue. (**C**) Magnitude (blue) and phase (orange) response of an arterial mock-up inflated to 75 mmHg. Phase is measured relative to stimulus. (**D**) Extending the measurements of magnitude (top) and phase (bottom) response across multiple mock-up pressures. Raw data points are plotted (circles) as well as their best-fit function provided by the Vector Fitting algorithm (*33*) (solid lines). Use of a fitting algorithm allows precise determination of peak location below the granularity of the frequency sweep (10 Hz steps).

Next, we verified the predicted leading order behavior of this resonance: an increase in pressure predicts an increase in resonant frequency. The mock-up was inflated to five internal pressures ranging from between 95-135 mmHg and an acoustic stimulus was swept across a wide range of frequencies while imaging the tubing with ultrasound (Fig. 2D). To determine the frequency of the resonant peak with precision beyond the 10 Hz granularity of our sweep, we applied the Vector Fitting algorithm, which identifies a best fit for the underlying frequency dynamics of a system (*33*). We then measured the resonant frequency of two mock-ups with different diameters over a range of internal pressures (60-150 mmHg) (Fig. 3). We observed that the resonant frequency increased as internal pressure was increased, in line with predictions from the physical model.

**Figure 3:**
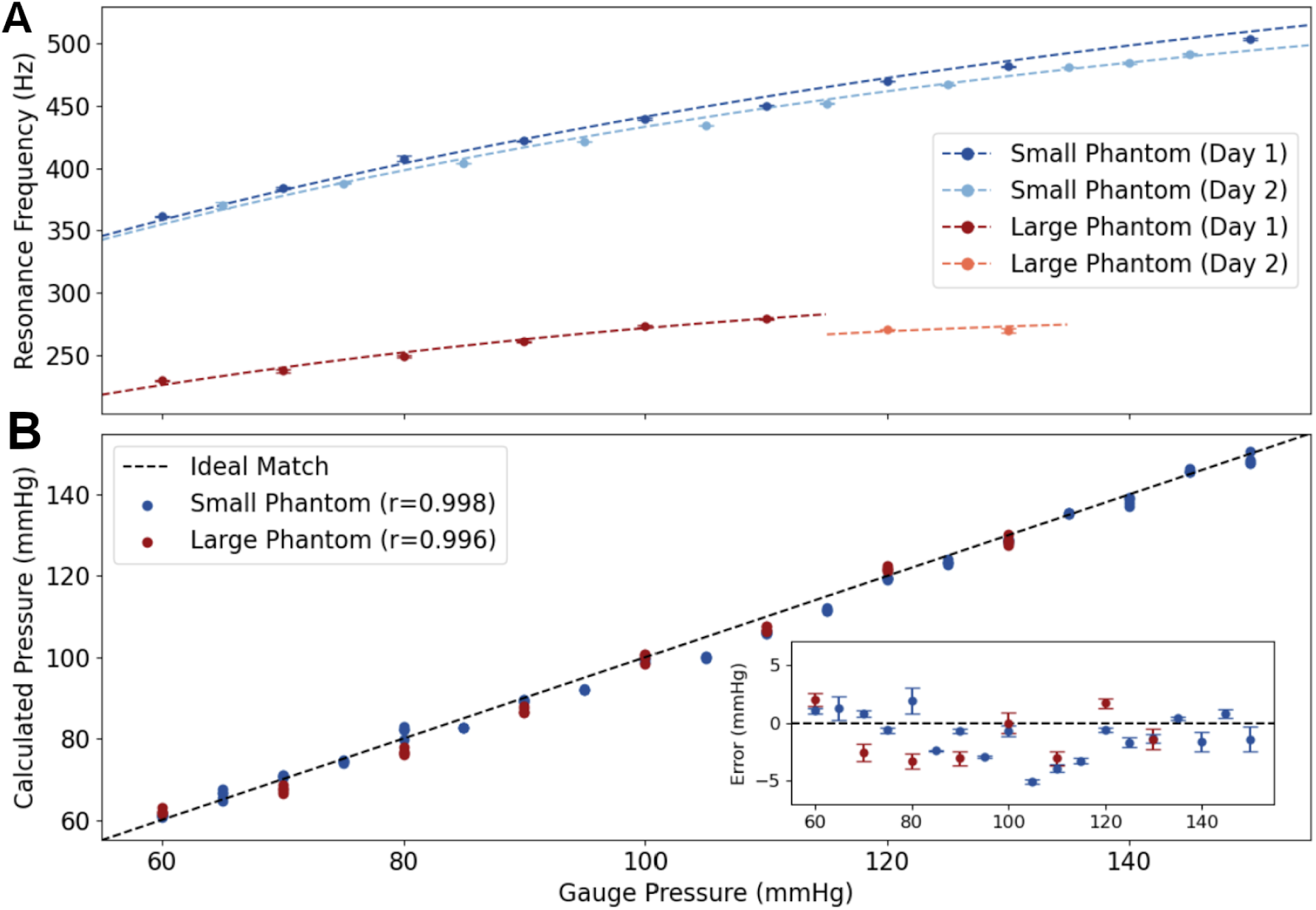
Comparison of predictions from the physical model and in vitro measurements. (**A**) Resonant frequency as a function of pressure, measured across two mock-ups with different radii. Blue and red markers indicate experimental data for small and large tubing, respectively. Dashed lines indicate predictions from the physical model. Data are separated by day as the mock-up was stored pressurized overnight, resulting in the next day’s experiments having a different zero-pressure diameter over the two different experiments. (**B**) Correlation between prediction from the physical model using VRR and imaging data versus the measured (gauge) pressure in the mock-up. Bottom right inset shows the deviations of the model pressure from the measured pressure in the mock-up. Markers and error bars denote mean and standard deviation of repeated trials (N=5 per data point).

We applied Equation 1 to predict pressure inside the mock-up based on our ultrasound-derived observables. Radius and thickness of the mock-up were measured at each pressure by analyzing the ultrasound imagery. The elastic modulus of the mock-up’s wall was computed by observing change in the pressure estimate versus the change in radius; this result was found to be in good agreement with independent tensometer measurements. Fig. 3A contains several pressure-frequency curves (different curve colors), demonstrating that the model correctly captures the effects of resonant frequency at different arterial dimensions. Fig. 3B depicts pressure as calculated by Equation 1 versus the gauge pressure; the inset plot shows the error between the two. Across all measurements the mean error between the values calculated via the physics model and the measured gauge pressure was -1.09 mmHg with a standard deviation of 1.98 mmHg.

### Validation of physical model in humans

While mock-ups allowed us to validate our model in a controlled setting, human arteries and physiology are considerably more complex. BP in the human body is far from constant, rapidly fluctuating from its peak at systole (systolic blood pressure, SBP) to the diastolic minimum (diastolic blood pressure, DBP) over a single heartbeat. Additionally, artery walls are not simple elastic membranes free-floating in a homogenous medium; they are made up of multiple anisotropic layers and are embedded in other tissues. To determine whether arterial resonance persists *in vivo*, we applied this methodology on the carotid artery in the neck.

A representative frequency response from a human carotid artery using acoustic stimulation is shown in Fig. 4A-D. Not only did the expected resonant behavior exist *in vivo*, but we were also able to resolve variations in this behavior over the course of the cardiac cycle. Arterial dimensions were estimated from feature detection and tracking in the B-mode imagery (Fig. S3). To reduce noise, raw radius and wall velocity estimates were combined using a linear Kalman filter to produce more consistent radius measurements. Raw pressure values were calculated at a rate of 200 Hz and then smoothed with a 20 Hz lowpass filter to produce final outputs. Acoustic stimulus was provided by a repeating, multi-frequency waveform with a 50 millisecond period (equivalent to 20 Hz).

**Figure 4:**
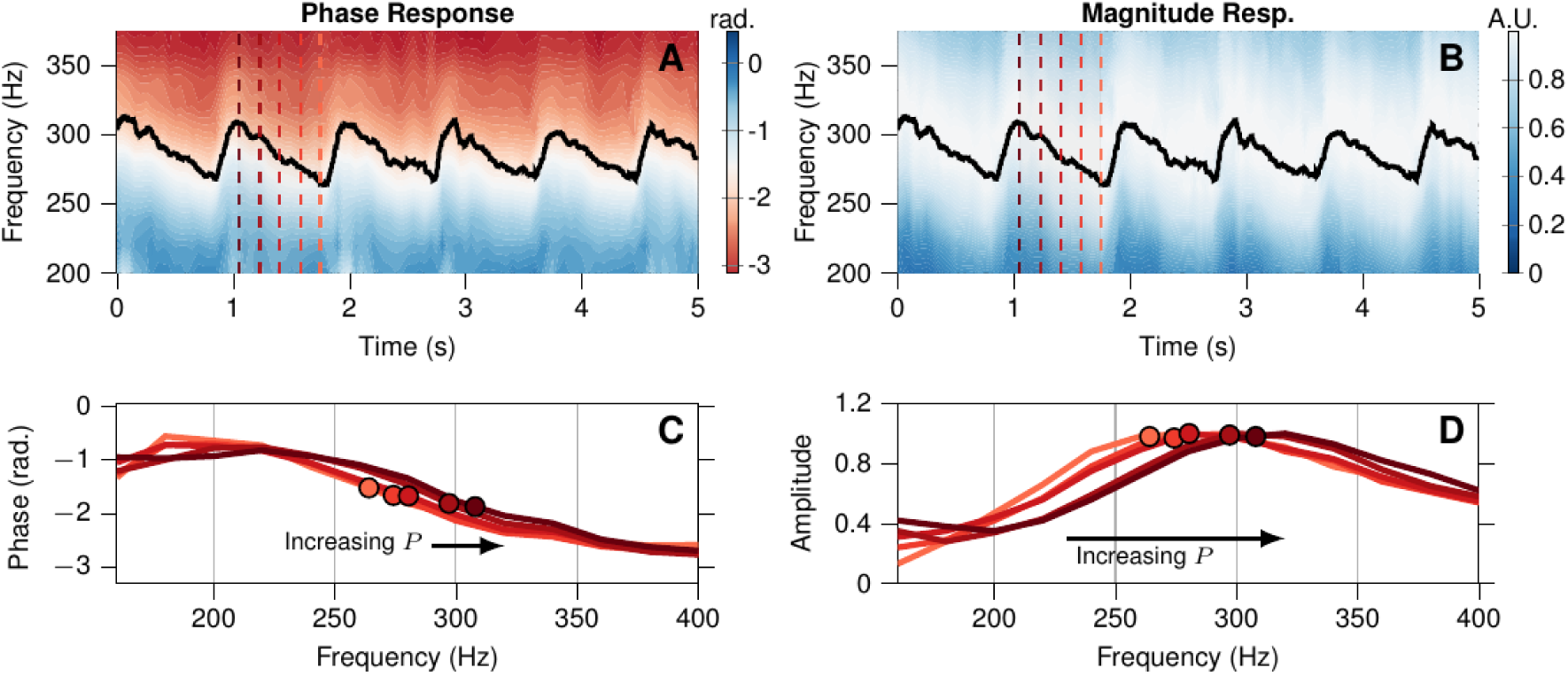
Excerpts of *in vivo* VRR from external audio-frequency stimulation data. (**A**) and (**B**) show contour maps of the phase and magnitude response, respectively, as a function of frequency over a 5-second window in a single subject. Solid black lines in these panels indicate the fitted resonant frequency. (**C**) and (**D**) are magnitude and phase response plots corresponding to time instances marked with the dashed red lines in panels A and B, with darker shades of red corresponding to higher pressure.

We successfully applied the method to the carotid artery (Fig. 5A, Movie S1) as well as several peripheral sites: the axillary (Fig. 5C), brachial (Fig. 5E) and femoral (Fig. 5G). VRR was stimulated and observed across all four artery sites; the resonant frequencies in Fig. 5B, D, F, and H (for the carotid, axillary, brachial, and femoral, respectively) vary throughout the cardiac cycle synchronously with the corresponding arterial radius measurements. At all four sites, the method produced blood pressure measurements in a single subject that were broadly in line with those obtained from an oscillometric cuff. Consistent with the physical model, the narrower cross-section of the brachial artery induces higher resonance frequencies (450-550 Hz) compared to those in the larger femoral and carotid arteries (*∼*270-350 Hz). For all measurements, the fitted resonant frequency was found to increase with the BP during systole, and then decrease again with diastole, synchronous to pulsation in arterial radius (Fig. 5B, D, F, and H) and consistent with the model.

**Figure 5:**
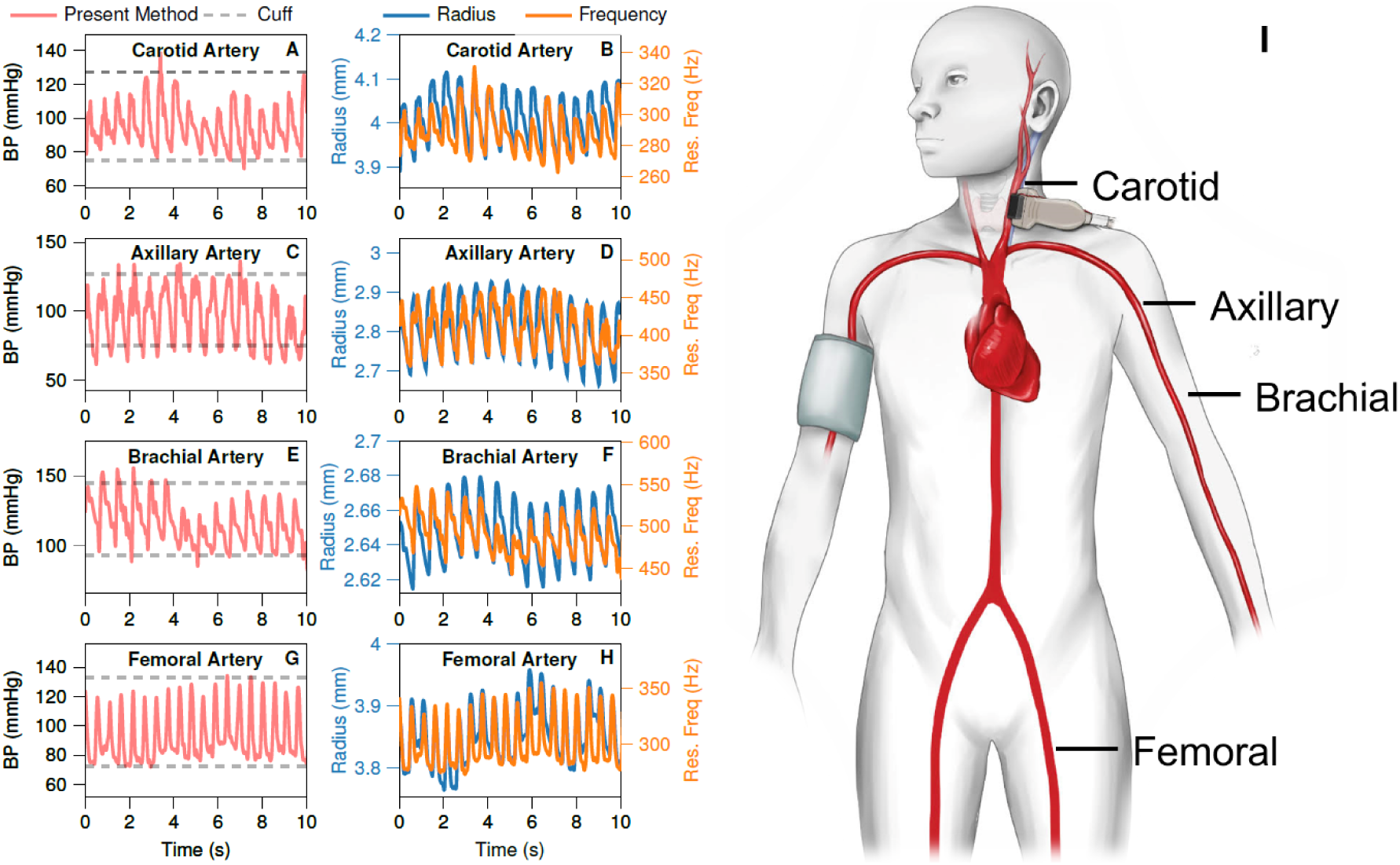
Comparison of predictions from the physical model and *in vitro* measurements. Measurement in carotid, femoral, and brachial arteries in one subject. (**A**), (**C**), (**E**), (**G**) Blood pressure waveforms (red) taken from four artery locations (carotid, axillary, brachial, and femoral, respectively) in a single subject compared with SBP and DBP obtained from an oscillometric cuff on the brachial artery (grey dashed lines). (**B**), (**D**), (**F**), (**H**) Measurements of arterial radius (blue) and resonant frequency (orange) corresponding to the BP waveforms in panels A, C, E, and G. (**I**) Diagram indicating the relative location of each artery. Note that because the BP cuff can only measure SBP and DBP intermittently, the values are extrapolated as constant values.

Additional preliminary testing was conducted on the carotid arteries of six human subjects, revealing similar frequency responses to the audio stimulus. We find that the new method successfully captures complete blood pressure waveforms in all six subjects, across genders and a small set of ages (Table S3). Fig. 6 shows continuous blood pressure waveforms in five second windows from all six subjects. Comparison of an intermittent BP cuff measurement to a continuous measurement is likely to have greater error due to respiratory variation (Fig. 6G). Critically, unlike the BP cuff, this method is not constrained to just measurements of systolic or diastolic pressure.

**Figure 6:**
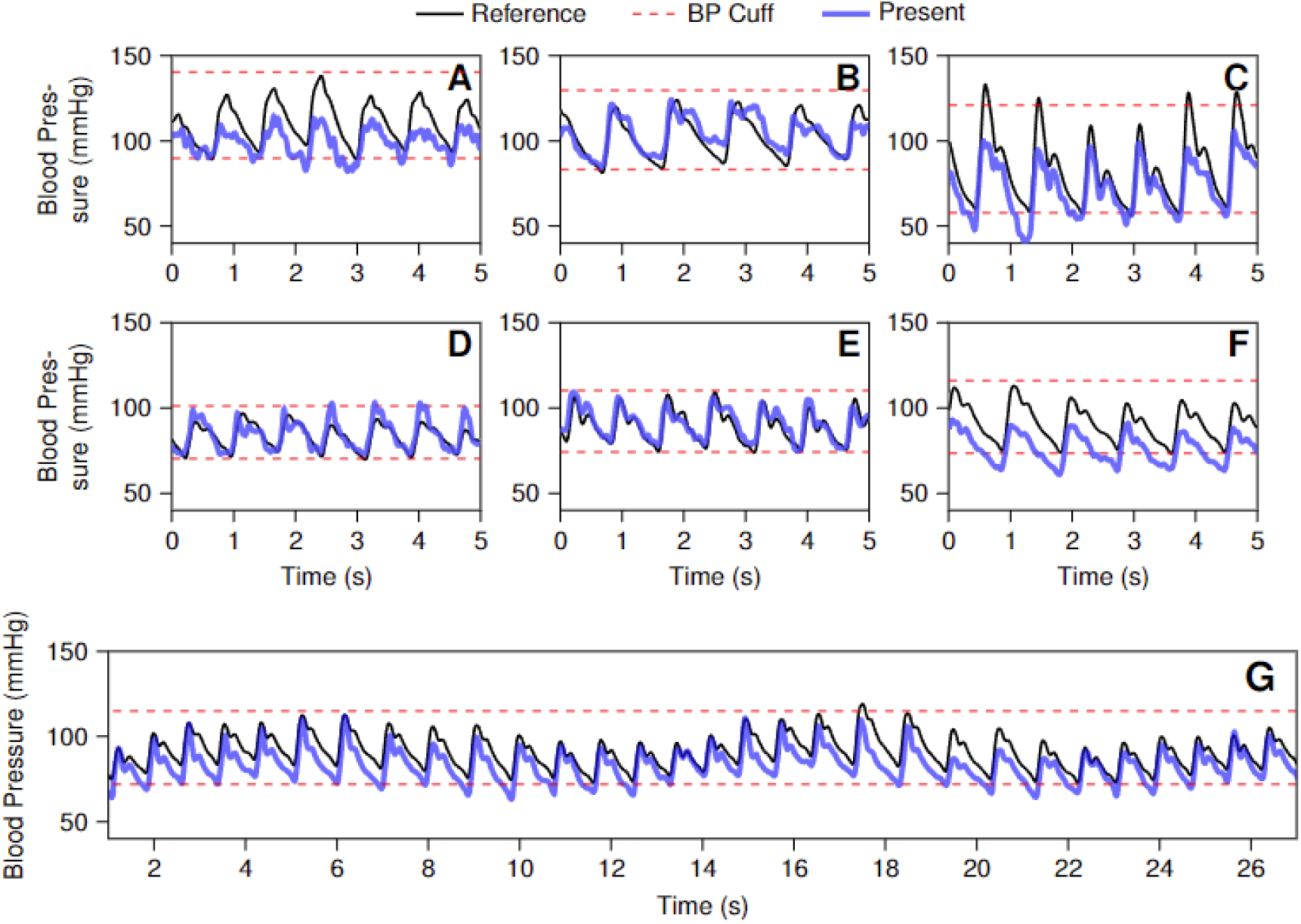
Representative waveforms of carotid pressure. Excerpts of *in vivo* data from the six subjects low-pass filtered at 25 Hz. Panels **A**-**F** correspond to excerpts from Subjects A-F, respectively. Each panel shows 5 seconds of pressure waveforms calculated by the present method, shown in blue, along with the reference systolic and diastolic pressures from the BP cuff, shown in red dashed lines. Panel **G** shows a full 30 second time course of blood pressure measurements, and the slow oscillatory behavior underlying the trace is consistent with the variation in blood pressure seen when a subject (subject F) breathes in and out (*∼* 2 breaths in the 30 second window). For comparison, the extrapolated pressure waveform pressure predicted by (*36*) is shown in solid black. Note that because the BP cuff can only measure SBP and DBP intermittently, the values are extrapolated as constant values.

Because the device was designed specifically for larger vessels (e.g., carotid), device contrast was not sufficient for consistent imaging of smaller vessels (e.g., brachial, radial). The comparison of carotid artery BP to a brachial BP cuff does not allow for comparison of SBP directly due to pulse amplification in peripheral vessels (*34, 35*). Consequently, the primary metric for comparison in this study is the DBP. The SBP values were significantly different (p*<*0.001), as expected for measurement on a central versus peripheral arterial site; however, the DBP was not significantly different (p=0.54) (Fig. 7).

**Figure 7:**
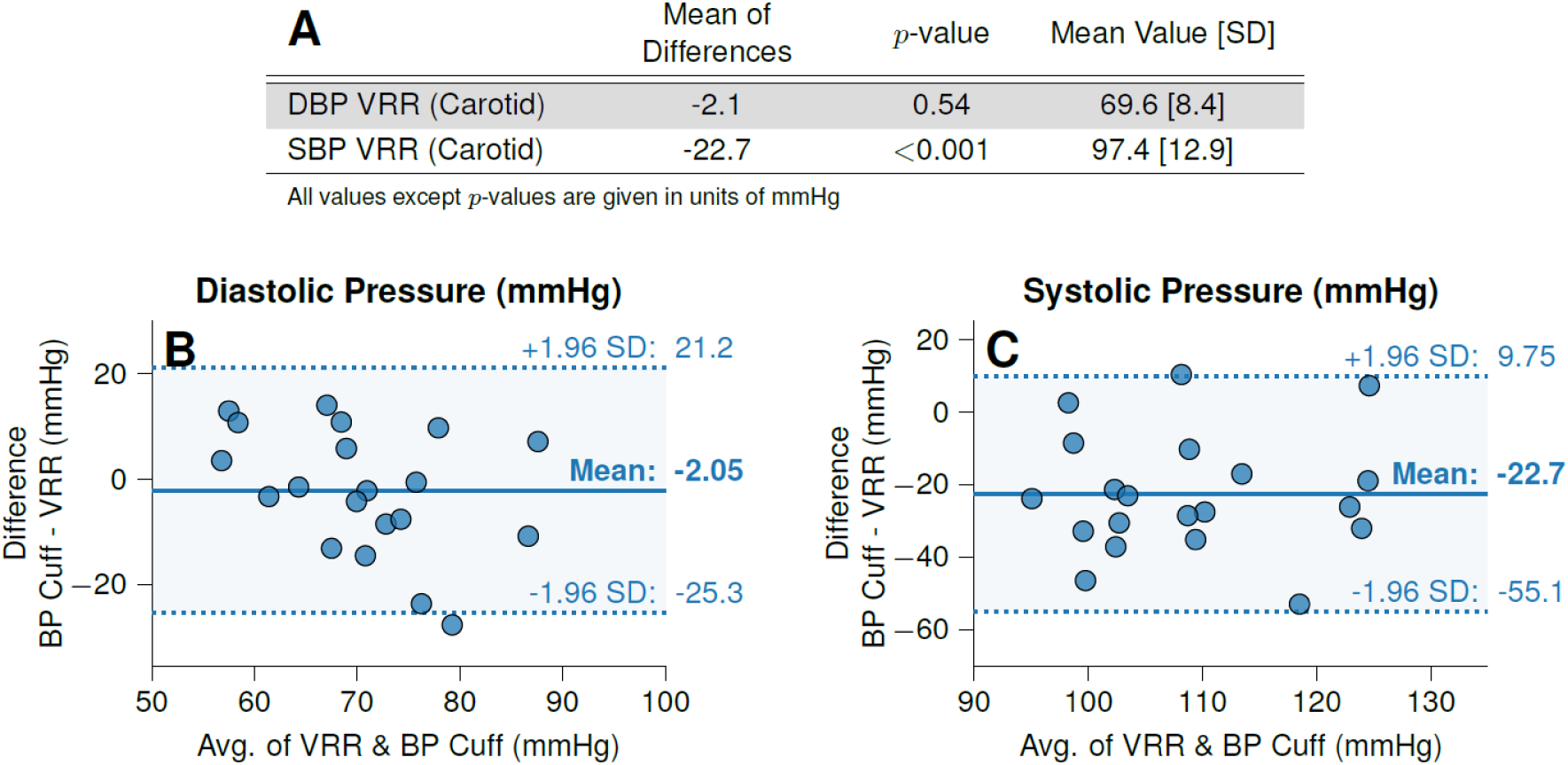
***In vivo* measurement Results for carotid VRR versus brachial cuff in six subjects. (A)** displays combined statistical values for VRR versus BP cuff. Bland-Altman plots of DBP **(B)** and SBP (**C**) for the carotid VRR to brachial cuff comparison.

## Discussion

In this study, we have demonstrated that VRR is a physical phenomenon predicted by classical mechanics and can be applied to the continuous measurement of BP in human arteries. Previous studies have demonstrated that ultrasound-based techniques can determine relative changes in BP, but passive ultrasound imaging alone is insufficient to establish an absolute baseline for pressure (*17–21, 36*). By perturbing the system with an acoustic stimulus, the method can obtain absolute BP without calibration to a reference device. Aside from measurements of SBP and DBP, the method is also able to obtain BP on a 50 millisecond basis, capturing the shape of the full BP waveform. This waveform can yield significant information that is not available from cuff measurements (*1–3, 37*).

We first validated the physical model *in vitro* using simple mock arteries where the material properties and dimensions could be controlled and verified. The model predicted pressures in agreement with the control pressure over a biologically relevant pressure range. Furthermore, we found that the model compensated for creep caused by stress-relaxation in both mock arter-ies, indicating that the model correctly accounts for changes in arterial radius versus resonant frequency.

We then successfully stimulated and detected VRR *in vivo* in four different arteries of a human subject and produced BP measurements consistent with those from a BP cuff. These results suggest that measuring continuous BP using this approach extends to other arteries and is not necessarily limited by size or location. A good qualitative agreement was observed between the device measurements (e.g., brachial) and those obtained from the BP cuff. Measurements obtained at the carotid artery return lower systolic values than the BP cuff, which is consistent with the carotid artery being located more centrally than the brachial artery. Second, our method is able to detect the modest low frequency variations (*∼*0.1 Hz) in BP associated with the subject’s respiration.

We focused on the carotid artery for the majority of our *in vivo* tests, as the carotid is a comparatively large and superficial vessel, making ultrasound measurements simpler to obtain, allowing us to evaluate the validity of the physical model more readily. Additionally, noninvasive measurements of central arterial pressure have significant scientific and clinical interest (*2, 38–42*) and are simply not possible with other popular methods such as volume clamping or photoplethysmography. Since the carotid is more central than arteries targeted by oscillometric cuffs (brachial artery in the upper arm) or arterial catheters (most often the radial artery in the wrist or femoral artery near the groin (*43*)), simple and noninvasive pressure measurements in the carotid could provide more insight for clinicians into the progression of cardiovascular disease.

A sensitivity analysis (Supplementary Text 2.3) was conducted in order to assess for potential sources of error, which identified frequency and radius as leading order terms with a scaling comparable to Equation 2. Although the stimulus is in the audio frequency range, transmission is coupled to the skin; thus, external audio stimuli (e.g., from ambient conversation) has no effect on extracting resonant frequency (Supplementary Text 2.4). Further, the Vector Fitting approach has been previously shown to be robust for identifying resonant frequency (*33*). In contrast, radius is likely a significant source of error in the present implementation due to the positioning (along the length of the artery instead of across) of the manually operated prototype device. However, minor disagreements with the cuff such as is seen in Fig. 6F (blue curve versus dashed lines) may actually indicate physiological changes in this individual’s cNIBP trace as a consequence of e.g., relaxation or respiration, demonstrating the power of the approach to show subtle variation in SBP and DBP in time not achievable by the cuff alone. Unlike the VRR approach, the reference method-based result plotted in the same figure (black curve) is forced into a potentially biased blood pressure range enforced by the cuff as a standard (*44, 45*). Future device implementations targeting more peripheral arteries (e.g. radial) will allow us to directly assess and validate similar BP fluctuations, ideally using the continuous gold standard arterial catheter as a reference.

A major strength of this method lies in the small number of observables required: arterial radius, thickness, and resonant frequency. The BP output is determined by utilizing these observables in real-time, without cuff information, prior training data, or any input of demographic information (e.g., BMI, age, gender, etc.). The findings also demonstrate that unlike other methods, VRR is not limited to a single peripheral artery but can be used in both central and peripheral arteries and is likely available to any artery visible by ultrasound. Because the application of this approach is straightforward and based on directly observable characteristics, integration of this method into wearable devices could one day make continuous BP waveform information widely and easily accessible.

## Supporting information

Movie S1, Animation_of_in_vivo_resonance.mp4

## Data Availability

The datasets generated and/or analyzed during the current study are available from the corresponding author on reasonable request.

## Acknowledgments

The authors would like to acknowledge Dr. Mikhail Shapiro and Bill Ling for early access to a Verasonics instrument for preliminary arterial mock-up imaging. The authors thank Laura Gleason for illustrations in figure panels. The authors would like to thank Lynne Rollins for her contributions. The authors would also like to thank Dr. Soheil Saadat for his discussions regarding statistical analysis.

## Authors’ contributions

A.R., A.B.R., D.Y., and Y.A. conceived of the method and worked through initial feasibility studies. R.J. designed the device, developed key algorithms & software, and performed initial analysis. S.D. helped with arterial mock-up data collection and experimental setup. A.B.R., W.P.D., M.K.F., A.C.R., and D.Y. performed data analysis. W.P.D., M.K.F., A.C.R., and D.Y. also performed algorithm development. A.B.R. and R.J. designed clinical studies. Clinical data were collected and analyzed by W.P.D., M.K.F., A.C.R., D.Y., R.J., and A.B.R. Y.A. helped with data analysis and model development. All authors contributed to the drafting and revision of the manuscript.

## Data and materials availability

All data are available in the manuscript or the supplementary materials.

## List of Supplementary Materials

• Figs. 1 to 7

• Materials and Methods

• Supplementary Text

• Figs. S1 to S3

• Tables S1 to S3

• Movie S1

## References

1. J. B. Mark, Direct Arterial Blood Pressure Monitoring: Normal Waveforms (Churchill Livingstone Inc., 1998), chap. 8, pp. 91–98.

2. D. Chemla, et al., Subendocardial viability ratio estimated by arterial tonometry: a critical evaluation in elderly hypertensive patients with increased aortic stiffness, Clinical and Experimental Pharmacology and Physiology 35, 909 (2008).

3. G. Moody, L. Lehman, Proceedings of Computers in Cardiology Conference (IEEE, 2009), vol. 36, pp. 541–544.

4. G. Nuttall, et al., Surgical and patient risk factors for severe arterial line complications in adults, Anesthesiology 124, 590 (2016).

5. J. E. Sharman, et al., Automated ‘oscillometric’ blood pressure measuring devices: how they work and what they measure, Journal of Human Hypertension (2022).

6. N. Kallioinen, A. Hill, M. S. Horswill, H. E. Ward, M. O. Watson, Sources of inaccuracy in the measurement of adult patients’ resting blood pressure in clinical settings: a systematic review, Journal of Hypertension 35, 421 (2017).

7. A. Bur, et al., Accuracy of oscillometric blood pressure measurement according to the relation between cuff size and upper-arm circumference in critically ill patients, Critical Care Medicine 28, 371 (2000).

8. A. J. Viera, K. Lingley, A. L. Hinderliter, Tolerability of the Oscar 2 ambulatory blood pressure monitor among research participants: a cross-sectional repeated measures study, BMC Medical Research Methodology 11 (2011).

9. P. Muntner, et al., Measurement of blood pressure in humans: A scientific statement from the American Heart Association, Hypertension 73, e35 (2019).

10. B. P. Imholz, W. Wieling, G. A. van Montfrans, K. H. Wesseling, Fifteen years experience with finger arterial pressure monitoring: assessment of the technology, Cardiovascular Research 38, 605 (1998).

11. P. Salvi, A. Grillo, G. Parati, Noninvasive estimation of central blood pressure and analysis of pulse waves by applanation tonometry, Hypertension Research 38, 646 (2015).

12. X. Quan, et al., Advances in non-invasive blood pressure monitoring, Sensors (Basel) 21 (2021).

13. P. Shaltis, A. Reisner, H. Asada, 2005 IEEE Engineering in Medicine and Biology 27th Annual Conference (IEEE, 2005), pp. 3970–3973.

14. B. Ibrahim, R. Jafari, Cuffless blood pressure monitoring from an array of wrist bioimpedance sensors using subject-specific regression models: Proof of concept, IEEE Transactions on Biomedical Circuits and Systems 13, 1723 (2019).

15. R. Mukkamala, et al., Toward ubiquitous blood pressure monitoring via pulse transit time: Theory and practice, IEEE Transactions on Biomedical Engineering 62, 1879 (2015).

16. C.-S. Kim, A. M. Carek, O. T. Inan, R. Mukkamala, J.-O. Hahn, Ballistocardiogram-based approach to cuff-less blood pressure monitoring: Proof-of-concept and potential challenges, IEEE Transactions on Biomedical Engineering 65, 2384 (2018).

17. B. W. Beulen, et al., Toward noninvasive blood pressure assessment in arteries by using ultrasound, Ultrasound in medicine and biology 37, 788 (2011).

18. J. Vappou, J. Luo, K. Okajima, M. D. Tullio, E. E. Konofagou, Non-invasive measurement of local pulse pressure by pulse wave-based ultrasound manometry (PWUM), Physiological Measurement 32 (2011).

19. J. Seo, H.-S. Lee, C. G. Sodini, Non-invasive evaluation of a carotid arterial pressure waveform using motion-tolerant ultrasound measurements during the valsalva maneuver, IEEE Journal of Biomedical and Health Informatics 25, 163 (2021).

20. C. Wang, et al., Monitoring of the central blood pressure waveform via a conformal ultrasonic device, Nature Biomedical Engineering 2, 687 (2018).

21. A. M. Zakrzewski, A. Y. Huang, R. Zubajlo, B. W. Anthony, Real-time blood pressure estimation from force-measured ultrasound, IEEE Transactions on Biomedical Engineering 65, 2405 (2018).

22. A. Avolio, et al., Challenges presented by cuffless measurement of blood pressure if adopted for diagnosis and treatment of hypertension, Pulse (2022).

23. Y. Kawano, Diurnal blood pressure variation and related behavioral factors, Hypertension Research 34, 281 (2011).

24. B. M. Deegan, et al., The effect of blood pressure calibrations and transcranial doppler signal loss on transfer function estimates of cerebral autoregulation, Medical Engineering & Physics 33, 553 (2011).

25. V. Volovici, N. L. Syn, A. Ercole, J. J. Zhao, N. Liu, Steps to avoid overuse and misuse of machine learning in clinical research, Nature Medicine 28, 1996 (2022).

26. R. N. Arnold, G. B. Warburton, Flexural vibrations of the walls of thin cylindrical shells having freely supported ends, Proceedings of the Royal Society A 197, 238 (1949).

27. D. J. Korteweg, Ueber die fortpflanzungsgeschwindigkeit des schalles in elastischen röhren, Annalen der Physik 241, 525 (1878).

28. J. C. Bramwell, A. V. Hill, The velocity of pulse wave in man, Proceedings of the Royal Society of London. Series B, Containing Papers of a Biological Character 93, 298 (1922).

29. Y. C. Fung, E. E. Sechler, A. Kaplan, On the vibration of thin cylindrical shells under internal pressure, Journal of the Aerospace Sciences 24, 650 (1957).

30. U. S. Lindholm, D. D. Kana, H. N. Abramson, Breathing vibrations of a circular cylindrical shell with an internal liquid, Journal of the Aerospace Sciences 29, 1052 (1962).

31. G. B. Warburton, Vibration of a cylindrical shell in an acoustic medium, Journal of Mechanical Engineering Science 3, 69 (1961).

32. P. A. Hasgall, et al., IT’IS database for thermal and electromagnetic parameters of biological tissues (2022). Doi:10.13099/VIP21000-04-1.

33. B. Gustavsen, A. Semlyen, Rational approximation of frequency domain responses by vector fitting, IEEE Transactions on Power Delivery 14, 1052 (1999).

34. A. L. Pauca, S. L. Wallenhaupt, N. D. Kon, W. Y. Tucker, Does radial artery pressure accurately reflect aortic pressure?, Chest 102, 1193 (1992).

35. M. K. Armstrong, et al., Brachial and radial systolic blood pressure are not the same, Hypertension 73, 1036 (2019).

36. J. M. Meinders, A. P. Hoeks, Simultaneous assessment of diameter and pressure waveforms in the carotid artery, Ultrasound in Medicine and Biology 30, 147 (2004).

37. P. Boutouyrie, et al., Association between local pulse pressure, mean blood pressure, and large-artery remodeling, Circulation 100, 1387 (1999).

38. A. Kollias, S. Lagou, M. E. Zeniodi, N. Boubouchairopoulou, G. S. Stergiou, Association of central versus brachial blood pressure with target-organ damage: Systematic review and meta-analysis, Hypertension 67, 183 (2016).

39. M. E. Safar, et al., Central pulse pressure and mortality in end-stage renal disease, Hypertension 39, 735 (2002).

40. L. Trudeau, Central blood pressure as an index of antihypertensive control: Determinants and potential value, Canadian Journal of Cardiology 30, S23 (2014).

41. E. Agabiti-Rosei, et al., Central blood pressure measurements and antihypertensive therapy: A consensus document, Hypertension 50, 154 (2007).

42. R. Mukkamala, D. Xu, Continuous and less invasive central hemodynamic monitoring by blood pressure waveform analysis, American Journal of Physiology-Heart and Circulatory Physiology 299, H584 (2010).

43. B. V. Scheer, A. Perel, U. J. Pfeiffer, Clinical review: Complications and risk factors of peripheral arterial catheters used for haemodynamic monitoring in anaesthesia and intensive care medicine, Critical Care 6 (2002).

44. D. S. Picone, et al., Accuracy of cuff-measured blood pressure: Systematic reviews and meta-analyses, Journal of the American College of Cardiology 70, 572 (2017).

45. B. G. Celler, et al., Are korotkoff sounds reliable markers for accurate estimation of systolic and diastolic pressure using brachial cuff sphygmomanometry?, IEEE Transactions on Biomedical Engineering 68, 3593 (2021).

