## Supplementary Materials for "Resonance sonomanometry for noninvasive, continuous monitoring of blood pressure"

#### Contents of this PDF

**1 Methods and Materials** **2**

**3 References for Materials and Methods and Supplementary Material** **25**

**4 Supplementary Figures and Tables** **32**

### Additional supplementary materials

- Movie S1, `Animation_of_in_vivo_resonance.mp4`:
  Visualization of resonance effect and calculated blood pressure in a human carotid over multiple heartbeats

### 1 Methods and Materials

#### 1.1 Physics Model

To determine the resonance versus pressure relationship, we model our system as a long, thin-walled cylindrical shell (i.e., $L \gg a$ and $a \gg h$). We assume that this shell has a uniform radius and thickness along its length and that it is surrounded by incompressible fluid. Such a system will support many natural modes of wall motion, each composed of a superposition of an axial component consisting of $m/2$ wavelengths along the length of the cylinder and a circumferential component consisting of $n$ wavelengths around the circumference of the cylinder, where $m$ is an integer equal to or greater than 1 and $n$ is an integer greater than 1. In cylindrical coordinates of axial location $z$ and azimuthal angle $\theta$, the radial displacement $w$ of each point on the surface at any given time $t$ can be expressed as a superposition of sinusoidal basis functions given by

$$w(z,\theta,t) = \Sigma_{m,n} A_{mn} \sin \frac{m\pi z}{L} \cos n\theta \cos 2\pi f t \tag{1}$$

for some scalar amplitude $A_{mn}$. The general solutions for the equations of motion of this system are quite complex for arbitrary $m$ and $n$. In a system with $L \gg a$, however, the contributions of the axial modes are greatly suppressed compared to the circumferential ones. Neglecting terms proportional to $a/L$, the resonant frequencies take the form of roots of a cubic polynomial (*1*):

$$0 = \kappa^3 - K_2\kappa^2 + K_1\kappa - K_0 \tag{2}$$

$$\kappa = \frac{4\pi^2 \rho_S a^2 (1-\nu^2)}{E} f^2 \tag{3}$$

$$K_0 = \frac{h^2(1-\nu)}{24a^2}\left(n^8 - 2n^6 + n^4\right) + \frac{Pa}{Eh}\alpha_1 \tag{4}$$

$$K_1 = \frac{1-\nu}{2}\left(n^4 + n^2\right) + \frac{h^2}{12a^2}\alpha_2 + \frac{Pa}{Eh}\alpha_3 \tag{5}$$

$$K_2 = 1 + \frac{3-\nu}{2}n^2 + \frac{h^2}{12a^2}\left(n^4 + n^2\right) + \frac{n^2}{1-\nu^2}\frac{Pa}{Eh} \tag{6}$$

$$\alpha_1 = \frac{1-\nu}{2}\left[\left(1 - \frac{h^2}{12a^2}\right)n^6 - n^4\right] \tag{7}$$

$$\alpha_2 = \frac{3-\nu}{2}n^6 - \frac{3+\nu}{2}n^4 + n^2 \tag{8}$$

$$\alpha_3 = \left(\frac{3-\nu}{2} - \frac{h^2}{12a^2}\right)n^4 - n^2 \tag{9}$$

Where $\kappa$, $K_i$, and $\alpha_i$ are dimensionless parameters.

In a damped system the lowest frequency resonant mode is generally the easiest to excite as first-order damping forces will increase with frequency for a given magnitude of displacement; thus, we focus our attention on the $n = 2$ mode. Finding the smallest real root of Equation 2 and converting from $\kappa$ back to $f$ with $n = 2$ yields

$$f_{vac}^2 = \frac{C_0 - \sqrt{C_0^2 - C_1}}{24\pi^2(1-\nu^2)ha^4\rho_S} \tag{10}$$

$$C_0 = 5Eh(3a^2 + h^2) + 12a^3P \tag{11}$$

$$C_1 = 12Ea^2h(36a^3P - 4ah^2P + 9Eh^3) \tag{12}$$

The frequency here is labeled $f_{vac}$ because it represents the natural frequency of the cylinder in a vacuum. However, in our system the shell both contains and is surrounded by fluid with non-zero mass, which adds to the effective inertia of the wall as it resonates. For a system with $L \gg a$ and $c \gg f_{vac}a$, this adds a simple proportional correction to find the natural frequency of the system with fluid (*2*):

$$\frac{f^2}{f_{vac}^2} = \left[1 + \frac{2n}{n^2+1}\frac{\rho_L}{\rho_S}\frac{a}{h}\right]^{-1} \tag{13}$$

$$f^2 = \frac{C_0 - \sqrt{C_0^2 - C_1}}{24\pi^2(1-\nu^2)a^3\delta} \tag{14}$$

$$\delta = ah\rho_S + \frac{4}{5}a^2\rho_L \tag{15}$$

Inverting Equation 14 to solve for $P$ yields

$$P = \frac{9\alpha^4 - 5\left(3\alpha + \alpha^3\right)D + 3D^2}{-4(9\alpha - \alpha^3) + 12D}E \tag{16}$$

$$\alpha = h/a \tag{17}$$

$$D = 4\pi^2(1-\nu^2)\frac{\rho a^2 f^2}{E} \tag{18}$$

$$\rho = \alpha\rho_S + \frac{4}{5}\rho_L \tag{19}$$

To gain some intuition about the behavior of this pressure equation we can linearize it for small $D$, yielding

$$P \sim \rho a^2 f^2 - O(\alpha^3 E) \tag{20}$$

Typical blood pressures range from 5-40kPa (40-300mmHg) (*3*), and typical carotid Young's moduli range from 100-1000 kPA with lower stiffness values at diastole (*4*). Thus, our calculation of pressure is dominated by the measurement of resonant frequency and radius in the regime of $\alpha \lesssim 0.1$.

#### 1.2 Measurement Technique

Calculating BP using Equation 16 requires knowledge of seven system parameters; radius, wall thickness, resonant frequency, wall Young's modulus, wall Poisson ratio, wall density, and fluid density. The first two parameters are routinely measured today using ultrasound imaging (*5*). In practice, thicknesses were calculated as $h = \overline{h} * \overline{a}/a$ (where overlines represent temporal averages), relying on the incompressibility of the arterial wall. Parameters 5-7 can be assumed to hold nearly constant (*6–8*). This leaves two parameters, resonant frequency and Young's modulus, which must be calculated by more involved means. Young's modulus is discussed in more detail below; the robustness of our estimate in discussed further in Supplementary Text 2.2. See Supplementary Text 2.3 for further analysis of the sensitivity of the BP estimate to the various parameters and constants for measurements conducted in the carotid artery.

##### 1.2.1 Resonant Frequency

Previous work has found that high-speed ultrasound imaging is capable of measuring the propagation of shear waves down the length of the arterial wall (*9*). We instead apply high-speed Doppler ultrasound imaging to measure the strength of wall motion around the arterial circumference under stimulation at various frequencies. While stimulus is applied via audio-frequency transducers, we simultaneously measure the velocities of the top and bottom arterial walls using a separate ultrasound transducer and standard Doppler velocimetry techniques (*10*). A lock-in amplifier (for static *in vitro* data) or Fourier transform (for time-varying *in vivo* data) is used on

$v_{diff}$ to extract magnitude and phase of the motion response at the stimulus frequency. The set of magnitude and phase values at all stimulus frequencies represents the frequency response of our system.

In order to extract a resonant frequency from this frequency response, we apply a method from electrical systems analysis known as Vector Fitting. In general, the frequency response $H(f)$ of any system can be approximated as a sum of rational functions:

$$H(f) = \Sigma_{m=1}^{N} \frac{r_m}{if - a_m} + d + fe \tag{21}$$

where $f$ is frequency, $a_m$ and $r_m$ are complex poles and residues respectively, and $d$ and $e$ are real linear offset parameters. In particular, a resonant system will have a complex conjugate pair of poles. Vector Fitting is an algorithm which uses iterative least-squares fitting to find an optimal set of values for $(r_m, a_m, d, e)$ which best match the observed frequency response of the system (*11*). The final fitted resonant frequency of the system is represented by the magnitude of our complex conjugate pair of poles. If resonance is not present, the Vector Fitting algorithm will return a set of purely real poles; this is additional confirmation that our data contain resonant behavior.

##### 1.2.2 Young's Modulus

Previous studies focused on calculating arterial Young's modulus *in vivo* have utilized the Moens-Kortweg (first term) and Bramwell-Hill (last term) equations, which are related but independent measures of pulse wave velocity down the length of the artery (*12*):

$$c = \sqrt{\frac{Eh}{\rho_L d}} = \sqrt{\frac{A}{\rho_L}\frac{dP}{dA}} \tag{22}$$

where $c$ is pulse wave velocity, $d$ is arterial diameter, and $A$ is arterial cross-sectional area. Rearranging the latter two terms and substituting $A = \pi a^2$ yields a useful equation for calculating

Young's modulus based on changes in pressure:

$$\frac{Eh}{2a} = \pi a^2 \frac{dP}{2\pi a da} \tag{23}$$

$$E = \frac{a^2}{h}\frac{dP}{da} \tag{24}$$

Prior studies calculated $dP/da$ using ultrasound imaging to measure $a$ and an external reference device such as a cuff or tonometer to measure $P$. We replace this external reference with pressure measurements generated by our device. This creates a recursive relationship, as these pressure measurements are themselves dependent on the value of $E$ we measure. This interdependency can be resolved using the Gauss-Seidel method. First, a physiologically reasonable value for $E$ is chosen as a starting point, and $P$ is calculated at all radii based on this value using Equation 16. These $P$ values are then used to calculate $E$ using Equation 23. By repeating these two steps, both $P$ and $E$ converge on a self-consistent set of values which satisfy both Equations 16 and 23 (e.g., fig. S2, table S2).

An important facet of this method of measuring $E$ is that it does not require $E$ to be constant across different radii. Instead, it provides instantaneous $E$ estimates at the same rate that pressure measurements are generated. Real arteries have been found to exhibit substantial variation in $E$ between systole and diastole (*4*), making dynamic $E$ measurement essential to obtaining accurate pulse pressures.

#### 1.3 Artery Mock-up Setup

To validate the present physics model, we used compliant, thin-walled rubber tubing, sourced from latex rubber balloons (Qualatex 160Q or similar) to simulate human vasculature. Tube-in-gelatin mock-ups are often used as an ultrasound teaching aid as they provide similar imaging properties to blood vessels embedded in tissue (*13, 14*). We substituted a water/psyllium fiber (Metamucil) mixture (*15*) for the gelatin/psyllium fiber mixture, as tubing can disbond from the gelatin as pressure (and thus also the tubing's radius) is changed, leading to air pockets and behavior disparate from real anatomy. Professional ultrasound tissue models are unsuitable for these experiments as they do not have similar elasticity in their vessel analogues; several vendors we investigated used rigid tubing, and all were flow-only (no pressure simulation). Two sizes of tubing were used (2.18mm radius, "small"; and 3.23mm radius, "large"). Wall thicknesses of each were 0.25mm and 0.28mm, with a density of 1.93 g/cm measured for both.

The small tubing was submerged to a depth of 2-3 cm in the water/psyllium fiber bath, modeling a depth similar to that of the human carotid artery. Psyllium fiber was used as a scattering medium to simulate surrounding tissue. The tubing was filled with water and inflated using a syringe to add pressure. Pressure was held constant for the course of a scan. Each scan consisted of a stimulus sweep from 200 to 600 Hz in 10 Hz steps with simultaneous measurement using the ultrasound transducer. Five scans were performed at each pressure, and pressure was swept from 60 to 150 mmHg (targeting a physiologically-relevant range) in 5 mmHg increments, for a total of 95 scans.

The experiment was repeated using the larger diameter tubing to confirm that the model holds across different vessel sizes. Pressures were swept from 60 to 150 mmHg in 10 mmHg increments. Above 130mmHg, we found that the tubing would start to rapidly expand in an uneven manner (where the radii of specific segments would expand unevenly, as normally seen during inflation). As this behavior is not seen in healthy arteries, we discarded data above 130mmHg, for a total of 40 scans.

#### 1.4 Measurement Apparatus

We constructed a custom ultrasound device to provide full insight into the signal processing chain and to allow accurate timing between the resonance-driving stimulus and ultrasound measurement. This device is comprised of two USRP N210 software-defined radios (National Instruments, Austin, TX), tied to custom transmit and receive electronics. The transmit chain consists of a threshold block, followed by a bipolar high-voltage pulse generator (a MAX4940, from Maxim Electronics, San Jose, CA). The receive chain consists of a matching network and low-noise amplifier with voltage-controlled gain (an AD8336 from Analog Devices, Wilmington, MA). The gain is set via software and provides time-gain compensation. These chains are isolated from one another via an automatic transmit-receive switch (a MD0100 from Microchip, Chandler, AZ). The device has one transmit chain and two receive chains, which are multiplexed to 32 ultrasound transducer pixels. In addition, the device has an amplified low-voltage output which feeds the audio-frequency stimulus transducers.

Downstream of the transmit/receive electronics, a commercial 6L3 linear ultrasound probe (Acuson, Mountain View, CA) provides conversion to ultrasound. Audio-frequency stimulus is provided by a set of audio-frequency bone conduction transducers (BC-10 from Ortofon, Nakskov, Denmark), mounted to the Acuson 6L3.

Capture bandwidth of the software-defined radios and associated electronics extends to 25 MHz, allowing for adequate localization and image reconstruction of pulses ranging from 3-6 MHz. Raw data is captured for post-processing by custom software.

#### 1.5 Arterial Mock-up Data Analysis

For each individual scan, radius was calculated from the average delay in echo timings between the top and bottom walls, and resonant frequency was calculated using the Vector Fitting method described above. Because the tubing walls were much thinner than those of *in vivo* arteries, thickness could not be determined accurately from our ultrasound imaging due to limited resolution. Instead, we used high-precision calipers to measure the unpressurized radius and thickness of the tubing ($a_0$ and $h_0$). Because the tubing was assumed to be incompressible ($\nu = 0.5$), a pressure-dependent thickness could be calculated as $h = h_0 * (r_0/r)$. These caliper measurements along with the weight of the tubing were also used to calculate its density.

The Young's modulus of the tubing was calculated by comparing radius and resonant frequency measurements across multiple scans at different pressures, as described above. We assumed that the tubing was linearly elastic, so a single value of $E$ was calculated which minimized the relative error in pressure as determined by Equations 16 and 23; this value came out to roughly 1.16 MPa. The balloon material was later analyzed with a tensile strength measurement instrument from Instron (Norwood, MA). This test yielded an average stiffness of 1.10 MPa which held nearly constant across our strain range, validating both our calculated value and our linearity assumption.

Measured radii were adjusted for each scan based on this fixed $E$ value to generate agreement between these two equations. For the larger tubing, obtaining alignment with theory required adding $h/2$ to all radii; this would be explained if peak echoes from this system corresponded to the inner rather than average radius of the tube. The measured values for radius, thickness, resonant frequency, and stiffness were combined with prior values for wall density, fluid density, and wall Poisson ratio in Equation 16 to generate the final calculated pressure values shown in Fig. 3.

#### 1.6 Human Feasibility Studies: Design and Data Collection

##### 1.6.1 Study 1

This study was a prospective observational feasibility study evaluating the carotid, brachial, axillary, and femoral arteries in a single test subject compared to a BP cuff. For each scan, the subject's BP was monitored intermittently with an oscillometric cuff. This study was developed under guidelines for self-experimentation (*16*), and one of the authors both collected the data and served as the subject over several sessions in 2022 and 2023. The study was undertaken in order to evaluate the feasibility of data collection in multiple arteries of one subject. Data were analyzed to produce arterial pressure waveforms displayed in Fig. 5; no further analysis was conducted.

##### 1.6.2 Study 2

This study was a prospective observational, first-in-human feasibility study evaluating the carotid arteries in six test subjects (table S3) compared to a BP cuff. The research protocol was approved by the California Institute of Technology (protocol #: 19-0971). The aim of this study was to assess feasibility of collecting data in a single artery across multiple subjects for the sole purpose of demonstrating that resonance phenomena exist in humans. Study 2 evaluated the carotid artery in six volunteer test subjects in comparison to a BP cuff. Recruitment and data collection occurred during one session in October 2022. Data were collected with the BP cuff on the arm ipsilateral to the device placed on the carotid artery (IEEE Std 1708a™-2019 (*17*) and ISO 81060-2 (*18*)). Two measurements were taken with the BP cuff prior to initial data collection. Five one-minute measurements were taken with the device then two additional BP cuff measurements were taken. An additional five one-minute device measurements were taken and finally two additional BP cuff measurements were taken at the completion of data collection. Data were analyzed for SBP and DBP as well as arterial waveform presence and shape. Statistical analyses were completed using SigmaPlot 15.0 (Systat Software Inc) and Matlab 2022a (Mathworks). A power analysis was completed assuming alpha of 0.05 and power of 0.80 with an estimated effect size of 5mmHg. In a study using a t-test to compare two methods, a sample of 20 measurements can be used to determine a difference between the two datasets. Statistics were calculated according to universal standard ISO 81060-2.

It should be noted that, as an early feasibility study demonstrating proof-of-concept, strict adherence to regulatory standards was not intended, specifically in relation to subject numbers. This study was designed only to demonstrate that arterial resonance is observable in human arteries. Existing standards such as IEEE 1708 and ISO 81060-2 were not designed for cuffless, continuous, calibration-free devices and thus are not entirely appropriate for demonstration of this technology; however, we attempted to follow the static testing guidelines in IEEE 1708 and 81060-2 as closely as possible. ISO 81060-3 (*19*) had not yet been published at the time of data collection. It is likely a new set of procedures will need to be developed for future regulatory testing of this device.

#### 1.7 Data Processing Pipeline for Blood Pressure Calculation

To calculate BP, we extracted the necessary physical measurements required by the model (i.e., radius, resonance frequency, and thickness) from the ultrasound imaging. Using a custom data processing pipeline, we were able to extract the relevant measurements from the raw ultrasound data and produce a BP estimate. A detailed outline of the data processing pipeline is shown in fig. S1, with the key steps briefly summarized below. Using ultrasound imaging at a frame rate of $\sim 300$ Hz, we visualized the pulsatile artery walls in each subject and segmented them to extract instantaneous radius and thickness measurements (fig. S3, Movie S1). Using simultaneously captured Doppler ultrasound data, we also measured arterial wall velocities generated by our stimulus and used this response to identify the arterial resonant frequency at a rate of 200 Hz (Movie S1). User-specified indicators for the locations of the artery walls were also provided to aid in distinguishing the location of arterial walls within the image from other vasculature. The instantaneous elastic modulus was estimated continuously to account for the nonlinear elasticity of the arterial wall (Materials and Methods, Supplementary Text 2.2). A Kalman filter was used to combine wall velocity and image-derived radius & thickness measurements, providing a better estimate of both radius and thickness. These data were then fed into our stiffness calculator and the BP formula (Equation 1 in the main text). The resulting BP estimates were low-passed to remove unphysical high-frequency noise, and were then screened using a set of quality control (QC) criteria to remove data contaminated by artifacts stemming from the manual operation of the device, such as excessive motion of either the probe or subject.

#### 1.8 Quality Control Criteria and Data Exclusion

BP measurements obtained from the test device were processed using standard methods from the literature that are consistent with those employed by vital sign monitors (*20*). First, measurements were passed through an interquartile range filter and a 12 Hz lowpass filter. To convert continuous measurements into clinically relevant metrics of diastolic blood pressure (DBP) and systolic blood pressure (SBP), data were divided into non-overlapping time windows with a length of six seconds, rounded to the nearest heartbeat interval. For each window, DBP and SBP were calculated as the average of peak minima and maxima, respectively.

Because a manual placement of the probe is required for the current implementation of the device, measurements are highly sensitive to motion-induced operator error, including shifts due to operator fatigue, as well as test subject movement. As a result, signal loss was a regular occurrence and these data were deemed unusable and excluded from the final analysis.

#### 1.9 Calibrated Pressure Waveform from Arterial Radius

A reference for the arterial blood pressure at the carotid artery was calculated in Fig. 6 using the calibrated exponential function proposed by Meinders (2004). This empirically-derived relationship provides a method to obtain localized blood pressure from arterial radius waveforms by first calibrating the fitting parameters in the equation to a separate blood pressure reference, such as a brachial sphygmomanometer, when measurements from an arterial catheter are unavailable. This method has been employed in numerous previous studies as a means of extrapolating instantaneous blood pressure information from arterial dimensions obtained with ultrasound imaging (*12, 22–25*). Assuming that the target artery has a circular interior crosssection and exhibits minimal hysteresis, the pressure-radius relationship is given by

$$P(r) = P_{dia} \exp\left[\alpha\left(\frac{r^2}{r_{dia}^2} - 1\right)\right] \tag{25}$$

where $\alpha$ is an artery-specific stiffness coefficient given by

$$\alpha = \frac{r_{dia}^2}{r_{sys}^2 - r_{dia}^2} \ln\left[\frac{P_{sys}}{P_{dia}}\right]. \tag{26}$$

Here, $P$ is the arterial pressure, $r$ is the arterial radius, and the $_{dia}$ and $_{sys}$ subscripts indicate the diastolic and systolic values of the given parameters.

While $r$ and $r_{dia}$ can be measured directly with ultrasound at the target location, $P_{dia}$, $P_{sys}$, and, subsequently, $\alpha$ are not known *a priori* from passive ultrasound imaging and must be informed by an external blood pressure reference, in this case, a blood pressure cuff (Philips Intellivue MP70).

Because the reference blood pressure measurement might not be collected with the measurements of the arterial dimensions, as is the case of the present carotid artery measurements, Meinders (2004) suggested a correction whereby $\alpha$ is iteratively updated until the mean blood pressure obtained from the exponential fit matches that obtained from the reference. This approach operates under the assumption that the diastolic and mean blood pressures do not vary significantly in the various arteries, unlike systolic blood pressure (*26*). In the present study, $\alpha$ is first estimated using Equation 26 and then updated using

$$\alpha_{i+1} = \left(\left(\frac{\overline{P_{ref}}}{\frac{1}{T}\int_{t_0}^{t_0+T} P(r(t))dt} - 1\right)\beta + 1\right)\alpha_i \tag{27}$$

where $\overline{P_{ref}}$ is the reference mean blood pressure, and $\beta$ is a dimensionless over-relaxation factor to accelerate convergence. In instances where the mean blood pressure is not available, as can often be the case for brachial sphygmomanometer, $\overline{P_{ref}}$ can be estimated using the formula proposed by Meaney (2000):

$$\overline{P_{ref}} = P_{dia} + 0.412(P_{sys} - P_{dia}). \tag{28}$$

Iterative updating of $\alpha$ is conducted until the integrated estimate for mean blood pressure is within $\pm 0.01$ mmHg of $\overline{P_{ref}}$, which typically occurs with $\mathcal{O}(10)$ iterations.

### 2 Supplementary Text

#### 2.1 Further Detail of Existing Continuous Noninvasive BP Methods Versus Present Method

The performance and convenience gap between the catheter and cuff has inspired a number of methods aimed at enabling continuous, noninvasive BP measurements (*28*). We list them briefly to highlight the large body of work that has attempted to bridge this gap: finger cuffs (volume clamp method) (*29*), photoplethysmography (PPG) correlation (*30*), pulse transit time analysis (*31*), ballistocardiography (*32*), tonometry (*33*), capacitance measurement (*34*), electrical impedance measurement (*35*), radar measurement (*36*), and lastly, ultrasound measurement (*12, 22, 24, 25*) (table S1 for further details and comparisons). Noninvasive methods that do not require calibration are few and have inherent limitations such as periodic data blackouts (*37*) or the requirement of "black-box" machine learning techniques that may overfit to the demographics of the underlying dataset (*38*). We also note that these methods may not extend to all patients; for example, finger cuffs may be inaccurate in patients with peripheral vasoconstriction or hypotension (*39*).

Ultrasound-based methods, in particular, present several advantages compared to many of the previously listed alternatives. These methods can image deep arteries (unlike PPG or tonometry) and are unaffected by factors such as skin tone or body hair (unlike most optical methods). Additionally, ultrasound is already widely used in various diagnostic applications due to its safety and convenience. Numerous groups have proposed several methods for monitoring BP via ultrasound imagery by extracting parameters such as arterial wall diameter (*21, 22*) and blood flow velocity (*12, 24, 25*). However, these previous methods are limited to measurements of pulse pressure (i.e., the difference between systolic and diastolic pressure) and cannot establish an artery's baseline pressure without calibration to an externally obtained BP reference, such as a BP cuff. Because artery physiology (e.g., muscle tone or amount of vasoconstriction) changes over time, any method that relies on calibration also requires periodic recalibration to compensate for those changes (*40*). The timeframe and conditions when a calibration point is valid may vary drastically depending on the time of day or the physiological state of an individual (*41*). This constraint significantly limits the usefulness of these methods in dynamic conditions such as those found in critically ill patients, where vital signs can rapidly change, as well as in more common conditions such as during exercise.

Zakrzewski (2018) demonstrates measurements of absolute BP by combining ultrasound imaging, manually applied probe pressure, and a complex simulated tissue deformation model. This approach differs from other examined methods in a central way: rather than just passively imaging the artery, the distension of an artery is measured with respect to external stimulus (not dissimilar to the pressure applied by BP cuffs and finger cuffs). Though actively distending the imaged artery provides sufficient information to calculate the baseline diastolic pressure, the method still requires the use of a data-driven correction factor to obtain accurate results.

In contrast to these previous approaches, we present a new method for determining blood pressure within an artery (Fig. 1) that overcomes the shortcomings of prior ultrasound methods and requires no calibration or external reference. We start by treating a short section of artery as a hoop of elastic material, placed under tension by the pressure within. The core of this method is to then apply an acoustic stimulus (Fig. 1B) to the system, which causes a resonant response (Fig. 1C). Using ultrasound, we measure this resonance as well as the artery's dimensions and feed these values into a physical model; these data can be obtained from a single ultrasound transducer. Given these values, the physical model allows us to calculate the absolute pressure within the artery.

#### 2.2 Robustness of Young's Modulus Estimation

##### 2.2.1 Empirical Approach

An important consideration for our approach to calculating the circumferential Young's modulus is that arterial stiffness varies widely between subjects. For an arbitrary system of equations there is no guarantee that an iterative procedure, such as the one employed in this study, will always converge to a unique solution. We wish to show that for our particular system we do obtain robust convergence to a unique solution from any reasonable initial conditions. To do this, we performed an empirical analysis of convergence at 600 time steps randomly sampled from our *in vivo* carotid measurements (100 from each subject examined in Fig. 6). Previous studies have found that circumferential Young's modulus for the carotid artery varies from $0.1$ MPa to $1$ MPa in healthy adult subjects (*4*). To account for potential variations due to age or pathologies, we extended our analysis by a full order of magnitude in either direction, starting our iterative solver with seven initial values for Young's modulus ranging from $0.01$ MPa to $10$ MPa in geometric steps of $\sqrt{10}$. Gauss-Seidel iteration was performed for five steps from each starting value, and the seven final results for each of blood pressure ($P$) and Young's modulus ($E$) were compiled to compute coefficients of variation ($CV$) (defined as the standard deviation divided by mean) for each sampled time step.

Results for all six subjects are outlined in Table S2; the median $CV$ was less than $0.11\%$ for $E$ and less than $0.01\%$ for $P$, indicating robust convergence to a unique solution for any reasonable starting value of Young's modulus. Fig. S2 shows representative plots of how this iterative convergence looks in practice for the various initial values. There do exist initial values that cause the iterative method to diverge. These values consist of extreme initial estimates, such as $E < 0$ or $E > 100$ MPa, and unphysical inputs, such as $dP/da < 0$. However, this analysis indicates that convergence should be expected for physically reasonable starting points and parameters.

##### 2.2.2 Analytical Approach

The above empirical analysis shows that our iterative procedure robustly converges for a wide range of initial estimates for $E$ encompassing the physiologically relevant range, even if the starting point is orders of magnitude off from the final value. However, we do not need to rely on random initial values for $E$; instead, we can use an approximate solution to the system of equations and improve the accuracy of the starting value. The physical model consists of equations 16-19 and 24 from Materials and Methods (reproduced below as Supp. Equations 29-33), which represent a system of differential equations which we must solve in order to determine $P$ and $E$.

$$P = \frac{9\alpha^4 - 5\left(3\alpha + \alpha^3\right)D + 3D^2}{-4(9\alpha - \alpha^3) + 12D}E \tag{29}$$

$$\alpha = h/a \tag{30}$$

$$D = 4\pi^2(1-\nu^2)\frac{\rho a^2 f^2}{E} \tag{31}$$

$$\rho = \alpha\rho_S + \frac{4}{5}\rho_L \tag{32}$$

$$E = \frac{a}{\alpha}\frac{dP}{da} \tag{33}$$

Such systems may, in principle, generate a family of many different solutions, in which case our $P$ would not be uniquely determined. For our particular system, the solution $(P, E)$ which satisfies these equations at any given instant is uniquely determined. From an analytical standpoint, the full equation 29 is intractable; however, we can analyze its linearized version (equation 20 in Materials and Methods):

$$P \approx \frac{5}{3}\pi^2\left(1-\nu^2\right)\left(ah\rho_S + \frac{4}{5}a^2\rho_L\right)f^2 - \frac{h^3}{4a^3}E$$
$$= \frac{5}{3}\pi^2\left(1-\nu^2\right)\left(\gamma\rho_S + \frac{4}{5}a^2\rho_L\right)f^2 - \frac{\gamma^3}{4a^6}E \tag{34}$$
$$E = \frac{a^3}{\gamma}\frac{dP}{da} \tag{35}$$

We have replaced the product $ah$ with $\gamma$ because prior studies have found the arterial wall to be very nearly incompressible (*6, 7*). For a fixed length of the arterial wall, its cross-sectional area must remain constant even as pressure changes; thus, $\gamma$ is a constant independent of changes in $a$. To analyze this system, we make the common assumption that our physical system behaves smoothly as radius changes without sharp discontinuities in pressure, stiffness, or frequency. Take two consecutive measurements where the radius has changed by a small quantity $\epsilon$. These will generate a system of four equations:

$$P(a) = \frac{5}{3}\pi^2\left(1-\nu^2\right)\left(\gamma\rho_S + \frac{4}{5}a^2\rho_L\right)f^2 - \frac{\gamma^3}{4a^6}E(a)$$
$$P(a+\epsilon) = \frac{5}{3}\pi^2\left(1-\nu^2\right)\left(\gamma\rho_S + \frac{4}{5}(a+\epsilon)^2\rho_L\right)\left(f+\epsilon\frac{df}{da}\right)^2 - \frac{\gamma^3}{4(a+\epsilon)^6}E(a+\epsilon)$$
$$E(a) = \frac{a^3}{\gamma}\frac{P(a+\epsilon)-P(a)}{\epsilon}$$
$$E(a+\epsilon) = \frac{(a+\epsilon)^3}{\gamma}\frac{P(a+\epsilon)-P(a)}{\epsilon}$$

where $\frac{df}{da} = \frac{f(a+\epsilon)-f(a)}{\epsilon}$. Note that we have used our smoothness assumption to neglect terms of the order $\epsilon\frac{d^2f}{da^2}$, $\epsilon\frac{d^2P}{da^2}$, or $\epsilon\frac{d^2E}{da^2}$. Given that our device is able to measure $a$ and $f$ (and thus $\epsilon$ and $df/da$) at each time step, our system of four equations has only four unknowns. Rewriting in linear algebra form, we get

$$
\begin{bmatrix} C_1 \\ C_2 \\ 0 \\ 0 \end{bmatrix} = \begin{bmatrix} 1 & 0 & \frac{\gamma^3}{4a^6} & 0 \\ 0 & 1 & 0 & \frac{\gamma^3}{4(a+\epsilon)^6} \\ \frac{a^3}{\gamma\epsilon} & \frac{-a^3}{\gamma\epsilon} & 1 & 0 \\ \frac{(a+\epsilon)^3}{\gamma\epsilon} & \frac{-(a+\epsilon)^3}{\gamma\epsilon} & 0 & 1 \end{bmatrix} \begin{bmatrix} P(a) \\ P(a+\epsilon) \\ E(a) \\ E(a+\epsilon) \end{bmatrix}
$$

$$
C_1 = \frac{5}{3}\pi^2\left(1-\nu^2\right)\left(\gamma\rho_S + \frac{4}{5}a^2\rho_L\right)f^2
$$

$$
C_2 = \frac{5}{3}\pi^2\left(1-\nu^2\right)\left(\gamma\rho_S + \frac{4}{5}(a+\epsilon)^2\rho_L\right)\left(f+\epsilon\frac{df}{da}\right)^2
$$

Solving this system and applying the limit of $\epsilon \to 0$ to simplify yields the unique solution

$$
P(a) = \frac{5\pi^2 f\left(1-\nu^2\right)}{9}\frac{4\rho_L a^2\left(4a^4 f - 5\gamma^2 f - 2a\gamma^2\frac{df}{da}\right) + 5\rho_S\gamma\left(4a^4 f - 3\gamma^2 f - 2a\gamma^2\frac{df}{da}\right)}{4a^4 - 3\gamma^2}
$$

$$
E(a) = \frac{8\pi^2 a^7 f\left(1-\nu^2\right)}{3\gamma}\frac{4\rho_L a\left(f + a\frac{df}{da}\right) + 5\rho_S\gamma\frac{df}{da}}{4a^4 - 3\gamma^2}
$$

This solution is not exact as it was generated from a linearized version of our full equations; however, it should represent a close approximation of the true values of $E$ and $P$. It has been shown that for any twice continuously differentiable nonlinear system, Gauss-Seidel iteration is guaranteed to converge to the ideal solution given an initial guess reasonably close to this solution (*43*). Thus, by using our approximate analytical solution as a starting point we can have even greater confidence that numerically solving the nonlinear system through iteration will converge to the true solution. As shown in Fig. S2, our linearized estimate does indeed consistently come close to our final values for $P$ and $E$.

#### 2.3 Sensitivity Analysis of the Physics Model

Understanding the parameters to which the calculated blood pressure is most sensitive is critical for establishing the accuracy requirements of ultrasound-based imaging. We quantify this sensitivity through a propagation of error analysis in the equations of the physical model. Three key assumptions related to arterial geometry and mechanics underpin this analysis and the physical model of arterial resonance:

1. The artery can be modeled as a long cylinder of uniform wall thickness ($h$) and radius ($a$).

2. The audio stimulus operates in a perturbative fashion such that any nonlinear effects from the induced resonance can be neglected. Further damping effects due to the finite viscosity of the internal or external media or viscoelasticity of the wall itself can be modeled as a linear effect for the range of displacements induced by the stimulus.

3. The circumferential Young's Modulus of the arterial wall ($E$) behaves in a linearly elastic manner in response to the small radius perturbations induced by the stimulus. Such an assumption does not preclude changes in $E$ over the course of a cardiac cycle, only that changes in radius induced by the stimulus are small compared to variations in radius encountered over the course of a heartbeat.

By dimensional analysis, the internal blood pressure ($P$) within the artery can be expressed as a function of the geometric and material parameters as

$$P = g_1(a, h, f, \rho_L, \rho_S, E, \nu).$$

As before, $f$ is the natural frequency of circumferential waves along the arterial wall, $\nu$ is the arterial wall Poisson ratio, $\rho_S$ is arterial wall density, and $\rho_L$ density of fluid around the artery.

For human arteries, we further make two assumptions regarding the material properties of the artery and surrounding media:

1. $\rho_S$, $\rho_L$, and $\nu$ can be taken to be nearly constant parameters with known variability of a few percent across subjects (*8*).

2. The measurements of the different parameters are independent, and correlations in uncertainty between different parameters are assumed to be negligible.

The internal pressure at a given point in the cardiac cycle can be related to the parameters in a reference state (denoted with a 0 subscript) through

$$\begin{aligned} P - P_0 \approx (f - f_0) \left.\frac{\partial P}{\partial f}\right|_0 + (a - a_0) \left.\frac{\partial P}{\partial a}\right|_0 + (h - h_0) \left.\frac{\partial P}{\partial h}\right|_0 + (E - E_0) \left.\frac{\partial P}{\partial E}\right|_0 + \\ (\rho_S - \rho_{S0}) \left.\frac{\partial P}{\partial \rho_S}\right|_0 + (\rho_L - \rho_{L0}) \left.\frac{\partial P}{\partial \rho_L}\right|_0 + (\nu - \nu_0) \left.\frac{\partial P}{\partial \nu}\right|_0 \end{aligned} \tag{36}$$

Here, we use $\approx$ to indicate that we are neglecting higher-order terms in our analysis which are assumed to be small if our deviations from the $0$ state are sufficiently small. Taking the variance of each side of Equation 36, and letting $\sigma_i$ denote the standard deviation of parameter $i$ allows us to express the standard deviation of the internal pressure ($\sigma_P$) as a function of the variability of the other parameters through

$$\begin{aligned} \sigma_P^2 \approx \left( \left.\frac{\partial P}{\partial f}\right|_0 \sigma_f \right)^2 + \left( \left.\frac{\partial P}{\partial a}\right|_0 \sigma_a \right)^2 + \left( \left.\frac{\partial P}{\partial h}\right|_0 \sigma_h \right)^2 + \left( \left.\frac{\partial P}{\partial E}\right|_0 \sigma_E \right)^2 \\ + \left( \left.\frac{\partial P}{\partial \rho_S}\right|_0 \sigma_{\rho_S} \right)^2 + \left( \left.\frac{\partial P}{\partial \rho_L}\right|_0 \sigma_{\rho_L} \right)^2 + \left( \left.\frac{\partial P}{\partial \nu}\right|_0 \sigma_\nu \right)^2 \end{aligned} \tag{37}$$

Considering now our physical model allows us to quantify the relative sensitivity of the internal pressure to each parameter. Taking partial derivatives of Equation 16 with respect to each parameter and squaring each side gives the following relations for the leading order term of each derivative:

$$\left(\left.\frac{\partial P}{\partial f}\right|_0\right)^2 = \frac{P_0^2}{f_0^2}\left[4 + \frac{h_0}{a_0}\frac{5E_0}{(1-\nu_0^2)\pi^2\rho_{L0}a_0^2 f_0^2} + \mathcal{O}\left(\alpha^2\right)\right] \tag{38}$$

$$\left(\left.\frac{\partial P}{\partial a}\right|_0\right)^2 = \frac{P_0^2}{a_0^2}\left[4 + \frac{h_0}{a_0}\left(\frac{5\rho_{S0}}{\rho_{L0}} - \frac{15E_0}{2(1-\nu_0^2)\pi^2\rho_{L0}a_0^2 f_0^2}\right) + \mathcal{O}\left(\alpha^2\right)\right] \tag{39}$$

$$\left(\left.\frac{\partial P}{\partial h}\right|_0\right)^2 = \frac{P_0^2}{h_0^2}\left[\frac{h_0^2}{a_0^2}\left(\frac{5E_0}{8(1-\nu_0^2)\pi^2\rho_{L0}a_0^2 f_0^2} - \frac{5\rho_{S0}}{4\rho_{L0}}\right)^2 + \mathcal{O}\left(\alpha^3\right)\right] \tag{40}$$

$$\left(\left.\frac{\partial P}{\partial E}\right|_0\right)^2 = \frac{P_0^2}{E_0^2}\left[\frac{h_0^2}{a_0^2}\frac{25E_0^2}{64(1-\nu^2)^2\pi^4\rho_{L0}^2 a_0^4 f_0^4} + \mathcal{O}\left(\alpha^3\right)\right] \tag{41}$$

$$\left(\left.\frac{\partial P}{\partial \rho_S}\right|_0\right)^2 = \frac{P_0^2}{\rho_{S0}^2}\left[\frac{h_0^2}{a_0^2}\frac{25\rho_{S0}^2}{16\rho_{L0}^2} + \mathcal{O}\left(\alpha^3\right)\right] \tag{42}$$

$$\left(\left.\frac{\partial P}{\partial \rho_L}\right|_0\right)^2 = \frac{P_0^2}{\rho_{L0}^2}\left[1 - \frac{h_0}{a_0}\left(\frac{5\rho_{S0}}{2\rho_{L0}} - \frac{5E_0}{4(1-\nu_0^2)\pi^2\rho_{L0}a_0^2 f_0^2}\right) + \mathcal{O}\left(\alpha^2\right)\right] \tag{43}$$

$$\left(\left.\frac{\partial P}{\partial \nu}\right|_0\right)^2 = \frac{P_0^2}{\nu_0^2}\left[\frac{4\nu_0^2}{(1-\nu_0^2)^2} + \frac{h_0}{a_0}\frac{5\nu_0^4 E_0}{(1-\nu_0^2)^3\pi^2\rho_{L0}f_0^2 a_0^2} + \mathcal{O}\left(\alpha^2\right)\right] \tag{44}$$

Note that we have expanded each error term about small values of the parameter $\alpha = h/a$ to give intuition about the relative scale of the uncertainties, since $\alpha$ is assumed to be a relatively small parameter for our system. The terms $f$, $a$, $\rho_L$, and $\nu$ all have order-unity leading terms, indicating that they will be relatively important in contributing error. In contrast, the terms $h$, $E$, and $\rho_S$ have order-$\alpha^2$ leading terms, indicating that they will be relatively unimportant in the total error budget.

We can substitute representative values into the above equation to quantify how much each term contributes to the overall error budget. Assigning values to each parameter corresponding to a typical carotid artery gives: $a_0 = 4$ mm, $f_0 = 270$ Hz, $h_0 = 0.6$ mm, $E_0 = 0.385$ MPa, $\rho_S = 1102$ kg/m$^3$, $\rho_L = 1050$ kg/m$^3$, and $\nu = 0.5$.

$$\frac{\sigma_P}{P_0} \approx \left[5.2\left(\frac{\sigma_f}{f_0}\right)^2 + 5.1\left(\frac{\sigma_a}{a_0}\right)^2 + 0.0004\left(\frac{\sigma_h}{h_0}\right)^2 + 0.019\left(\frac{\sigma_E}{E_0}\right)^2 \right.$$
$$\left. +0.035\left(\frac{\sigma_{\rho_S}}{\rho_{S0}}\right)^2 + 0.90\left(\frac{\sigma_{\rho_L}}{\rho_{L0}}\right)^2 + 0.58\left(\frac{\sigma_\nu}{\nu_0}\right)^2\right]^{1/2} \quad (45)$$

In this instance, we can see that less than $10\%$ of error in thickness, stiffness, or arterial wall density will propagate into error in our final pressure estimation. Because stiffness is ultimately an inferred parameter, estimates of stiffness will only be as accurate as the underlying measurements. However, the sensitivity analysis reveals that the accuracy of our pressure estimates is relatively insensitive to uncertainty in the wall stiffness. The primary contributions to uncertainty come from frequency and radius, followed by fluid density and the Poisson ratio of the wall. Using the IT'IS database, we can approximate the uncertainty in our mass density value for blood ($\sigma_{\rho_L}$) and blood vessels using their values sampled from the literature. There, the standard deviation of blood densities was $\sigma_{\rho_L} = 17$ kg/m$^3$ and blood vessel density was $\sigma_{\rho_S} = 64$ kg/m$^3$. The combined uncertainty from just these two parameters corresponds to $< 2\%$ error in the blood pressure.

#### 2.4 Effect of Ambient Sound on the Method

The VRR method uses an audio-frequency stimulus (in range of 200-800 Hz) to obtain information about the artery's resonance behavior. Ambient sounds in this range are widely present, but we have not found this noise to be a factor in our data or represented in any of our analyses.

The insensitivity to ambient noise can be explained by performing a transmission/reflection calculation at the air-to-skin boundary. At an air-water interface, water is an almost perfect reflector of acoustic waves (*44*). Because tissue is mostly water, the acoustic impedance of skin ($\sim 1.6 \times 10^6$ kg m$^{-2}$ s$^{-1}$ per (*45*) and the IT'IS database (*8*)) and water ($\sim 1.48 \times 10^6$ kg m$^{-2}$ s$^{-1}$) are comparable, which would lead to the same near-perfect reflector effect for acoustic waves. Thus, it is nearly impossible for ambient noise from non-contact sources to penetrate into the tissue with enough intensity and coherence to interfere with the acoustic input from the direct contact transducers used in this method. In contrast, acoustic interference in the carotid could be spurred by vocal cord vibration when a subject speaks. During the study, we made sure that subjects did not speak during data acquisition to both prevent the artery from escaping the field of view and also to avoid this potential problem. Clinically, any patient who warranted monitoring of central (carotid) blood pressures, such as in an intensive care setting, would likely be sedated, negating this issue entirely.

### 3 References for Materials and Methods and Supplementary Material

1. Y. C. Fung, E. E. Sechler, A. Kaplan, On the vibration of thin cylindrical shells under internal pressure, *Journal of the Aerospace Sciences* **24**, 650 (1957).

2. G. B. Warburton, Vibration of a cylindrical shell in an acoustic medium, *Journal of Mechanical Engineering Science* **3**, 69 (1961).

3. J. A. Narloch, M. E. Brandstater, Influence of breathing technique on arterial blood pressure during heavy weight lifting, *Archives of Physical Medicine and Rehabilitation* **76**, 457 (1995).

4. T. Khamdaeng, J. Luo, J. Vappou, P. Terdtoon, E. E. Konofagou, Arterial stiffness identification of the human carotid artery using the stress-strain relationship in vivo, *Ultrasonics* **52**, 402 (2012).

5. V. Marque, H. van Essen, H. Struijker-Boudier, J. Atkinson, I. Lartaud-Idjouadiene, Determination of aortic elastic modulus by pulse wave velocity and wall tracking in a rat model of aortic stiffness, *Journal of Vascular Research* **38**, 546 (2001).

6. A. Karimi, T. Sera, S. Kudo, M. Navidbakhsh, Experimental verification of the healthy and atherosclerotic coronary arteries incompressibility via digital image correlation, *Artery Research* **16**, 1 (2016).

7. T. E. Carew, R. N. Vaishnav, D. J. Patel, Compressibility of the arterial wall, *Circulation Research* **23**, 61 (1968).

8. P. A. Hasgall, *et al.*, IT'IS database for thermal and electromagnetic parameters of biological tissues (2022). Doi:10.13099/VIP21000-04-1.

9. C. Papadacci, *et al.*, Non-invasive evaluation of aortic stiffness dependence with aortic blood pressure and internal radius by shear wave elastography and ultrafast imaging, *IRBM* **39**, 9 (2018).

10. W. R. Hedrick, D. L. Hykes, Autocorrelation detection in color doppler imaging: A review, *Journal of Diagnostic Medical Sonography* **11**, 16 (1995).

11. B. Gustavsen, A. Semlyen, Rational approximation of frequency domain responses by vector fitting, *IEEE Transactions on Power Delivery* **14**, 1052 (1999).

12. B. W. Beulen, *et al.*, Toward noninvasive blood pressure assessment in arteries by using ultrasound, *Ultrasound in medicine and biology* **37**, 788 (2011).

13. M. Earle, G. D. Portu, E. DeVos, Agar ultrasound phantoms for low-cost training without refrigeration, *African Journal of Emergency Medicine* **6**, 18 (2016).

14. C. Richardson, S. Bernard, V. A. Dinh, A cost-effective, gelatin-based phantom model for learning ultrasound-guided fine-needle aspiration procedures of the head and neck, *Journal of Ultrasound in Medicine* **34**, 1479 (2015).

15. R. O. Bude, R. S. Adler, An easily made, low-cost, tissue-like ultrasound phantom material, *Journal of Clinical Ultrasound* **23**, 271 (1995).

16. B. P. Hanley, W. Bains, G. Church, Review of scientific self-experimentation: Ethics history, regulation, scenarios, and views among ethics committees and prominent scientists, *Rejuvenation Research* **22**, 31 (2019).

17. Standards Committee, IEEE standard for wearable, cuffless, blood pressure measuring devices: Amendment 1, *Standard IEEE Std 1708a$^{TM}$-2019*, IEEE Engineering in Medicine and Biology Society, New York, NY (2019).

18. Non-invasive sphygmomanometers — Part 2: Clinical investigation of intermittent automated measurement type, *International Standard ISO 81060-2:2018(E)*, International Organization for Standardiation and International Electrotechnical Commission, Geneva, CH (2018).

19. Non-invasive sphygmomanometers — Part 3: Clinical investigation of intermittent automated measurement type, *International Standard ISO/FDIS 81060-3:2022(E)*, International Organization for Standardiation and International Electrotechnical Commission, Geneva, CH (2022).

20. W. Pasma, L. M. Peelen, S. van Buuren, W. A. van Klei, J. C. de Graaff, Artifact Processing Methods Influence on Intraoperative Hypotension Quantification and Outcome Effect Estimates, *Anesthesiology* **132**, 723 (2020).

21. J. M. Meinders, A. P. Hoeks, Simultaneous assessment of diameter and pressure waveforms in the carotid artery, *Ultrasound in Medicine and Biology* **30**, 147 (2004).

22. C. Wang, *et al.*, Monitoring of the central blood pressure waveform via a conformal ultrasonic device, *Nature Biomedical Engineering* **2**, 687 (2018).

23. C. Wang, *et al.*, Bioadhesive ultrasound for long-term continuous imaging of diverse organs, *Science* **377**, 517 (2022).

24. J. Seo, H.-S. Lee, C. G. Sodini, Non-invasive evaluation of a carotid arterial pressure waveform using motion-tolerant ultrasound measurements during the valsalva maneuver, *IEEE Journal of Biomedical and Health Informatics* **25**, 163 (2021).

25. J. Vappou, J. Luo, K. Okajima, M. D. Tullio, E. E. Konofagou, Non-invasive measurement of local pulse pressure by pulse wave-based ultrasound manometry (PWUM), *Physiological Measurement* **32** (2011).

26. W. W. Nichols, M. O'Rourke, C. Vlachopoulos, *McDonald's Blood Flow in Arteries : Theoretical, Experimental and Clinical Principles* (CRC Press, 2011).

27. E. Meaney, others., Formula and nomogram for the sphygmomanometric calculation of the mean arterial pressure, *Heart* **84**, 64 (2000).

28. P. Muntner, *et al.*, Measurement of blood pressure in humans: A scientific statement from the American Heart Association, *Hypertension* **73**, e35 (2019).

29. B. P. Imholz, W. Wieling, G. A. van Montfrans, K. H. Wesseling, Fifteen years experience with finger arterial pressure monitoring: assessment of the technology, *Cardiovascular Research* **38**, 605 (1998).

30. P. Shaltis, A. Reisner, H. Asada, *2005 IEEE Engineering in Medicine and Biology 27th Annual Conference* (IEEE, 2005), pp. 3970–3973.

31. R. Mukkamala, *et al.*, Toward ubiquitous blood pressure monitoring via pulse transit time: Theory and practice, *IEEE Transactions on Biomedical Engineering* **62**, 1879 (2015).

32. C.-S. Kim, A. M. Carek, O. T. Inan, R. Mukkamala, J.-O. Hahn, Ballistocardiogram-based approach to cuff-less blood pressure monitoring: Proof-of-concept and potential challenges, *IEEE Transactions on Biomedical Engineering* **65**, 2384 (2018).

33. P. Salvi, A. Grillo, G. Parati, Noninvasive estimation of central blood pressure and analysis of pulse waves by applanation tonometry, *Hypertension Research* **38**, 646 (2015).

34. X. Quan, *et al.*, Advances in non-invasive blood pressure monitoring, *Sensors (Basel)* **21** (2021).

35. B. Ibrahim, R. Jafari, Cuffless blood pressure monitoring from an array of wrist bio-impedance sensors using subject-specific regression models: Proof of concept, *IEEE Transactions on Biomedical Circuits and Systems* **13**, 1723 (2019).

36. M. P. Ebrahim, *et al.*, Blood pressure estimation using on-body continuous wave radar and photoplethysmogram in various posture and exercise conditions, *Scientific Reports* **9** (2019).

37. B. M. Deegan, *et al.*, The effect of blood pressure calibrations and transcranial doppler signal loss on transfer function estimates of cerebral autoregulation, *Medical Engineering & Physics* **33**, 553 (2011).

38. V. Volovici, N. L. Syn, A. Ercole, J. J. Zhao, N. Liu, Steps to avoid overuse and misuse of machine learning in clinical research, *Nature Medicine* **28**, 1996 (2022).

39. C. Ilies, *et al.*, Investigation of the agreement of a continuous non-invasive arterial pressure device in comparison with invasive radial artery measurement, *British Journal of Anaesthesia* **108**, 202 (2012).

40. A. Avolio, *et al.*, Challenges presented by cuffless measurement of blood pressure if adopted for diagnosis and treatment of hypertension, *Pulse* (2022).

41. Y. Kawano, Diurnal blood pressure variation and related behavioral factors, *Hypertension Research* **34**, 281 (2011).

42. A. M. Zakrzewski, A. Y. Huang, R. Zubajlo, B. W. Anthony, Real-time blood pressure estimation from force-measured ultrasound, *IEEE Transactions on Biomedical Engineering* **65**, 2405 (2018).

43. M. Vrahatis, G. Magoulas, V. Plagianakos, From linear to nonlinear iterative methods, *Applied Numerical Mathematics* **45**, 59 (2003).

44. O. A. Godin, Sound transmission through water-air interfaces: new insights into an old problem, *Contemporary Physics* **49**, 105 (2008).

45. C. Kuhn, F. Angehrn, O. Sonnabend, A. Voss, Impact of extracorporeal shock waves on the human skin with cellulite: A case study of an unique instance, *Clinical Interventions in Aging* **3**, 201 (2008).

46. B. Jana, K. Oswal, S. Mitra, G. Saha, S. Banerjee, Windkessel model-based cuffless blood pressure estimation using continuous wave doppler ultrasound system, *IEEE Sensors Journal* **20**, 9989 (2020).

47. J. Fortin, *et al.*, A novel art of continuous noninvasive blood pressure measurement, *Nature Communications* **12**, 1387 (2021).

48. J. Sola, *et al.*, Validation of the optical aktiia bracelet in different body positions for the persistent monitoring of blood pressure, *Scientific Reports* **11**, 20644 (2021).

49. K. Takazawa, H. Kobayashi, N. Shindo, N. Tanaka, A. Yamashina, Relationship between radial and central arterial pulse wave and evaluation of central aortic pressure using the radial arterial pulse wave, *Hypertension Research* **30**, 219 (2007).

50. C. Liao, O. Shay, E. Gomes, N. Bikhchandani, *2021 IEEE 17th International Conference on Wearable and Implantable Body Sensor Networks (BSN)* (2021), pp. 1–5.

### 4 Supplementary Figures and Tables

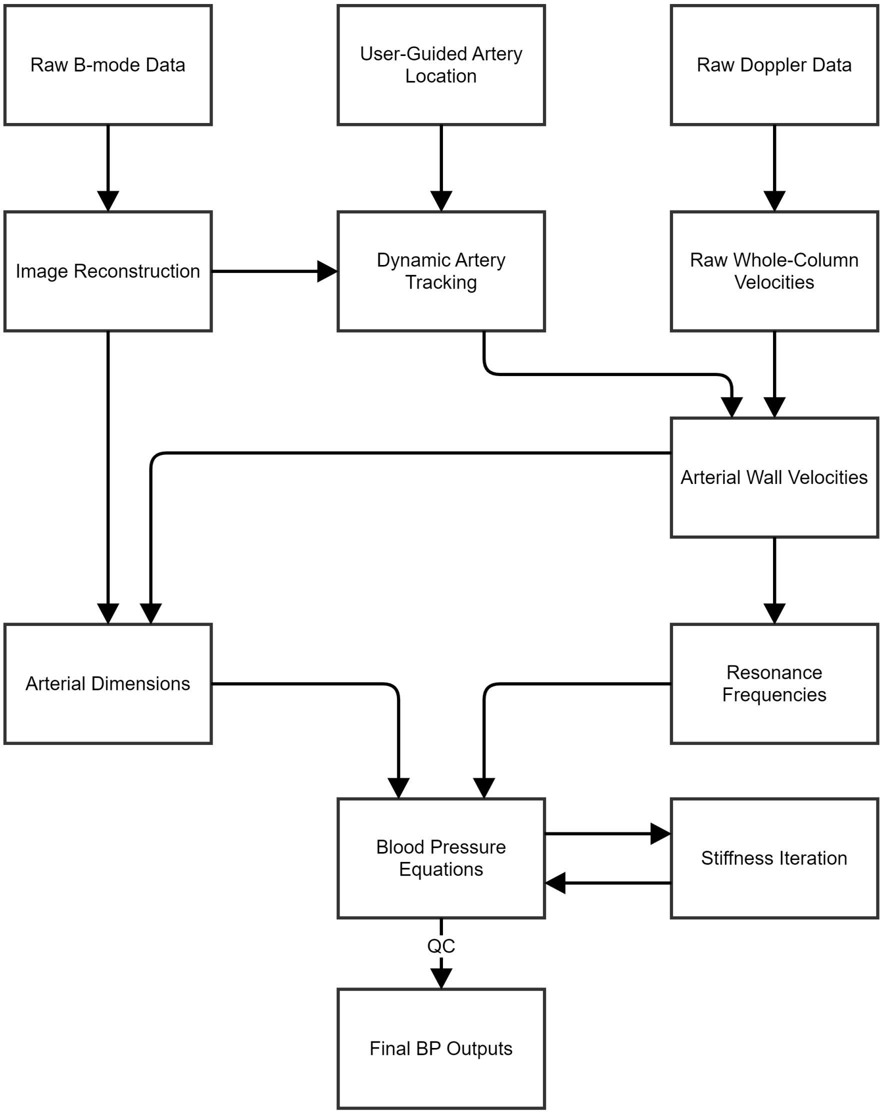

Figure S1: **Illustration of data processing pipeline.** Each box represents an input or algorithm; these produce the final data products seen in Figs. 4-7.

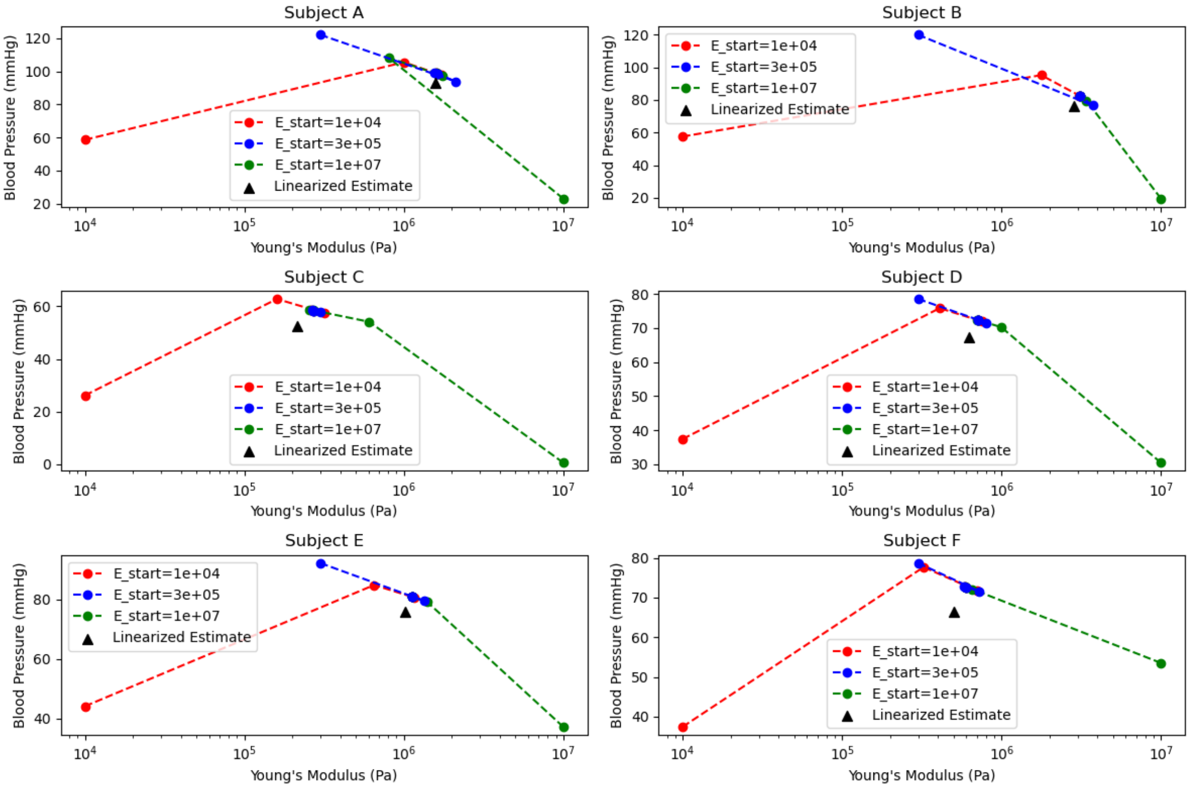

Figure S2: **An illustration of the convergence in $E$, viewed over several iterations.** Each panel represents one randomly chosen measurement point from each subject and shows our iteration from three out of the seven initial conditions; $E = 0.01$ MPa, $E = 0.3$ MPa, and $E = 10$ MPa. In every case, all three iterative traces quickly converge to the same location. Furthermore, in every case, this convergent location is close to what we would have estimated from our linearized solution.

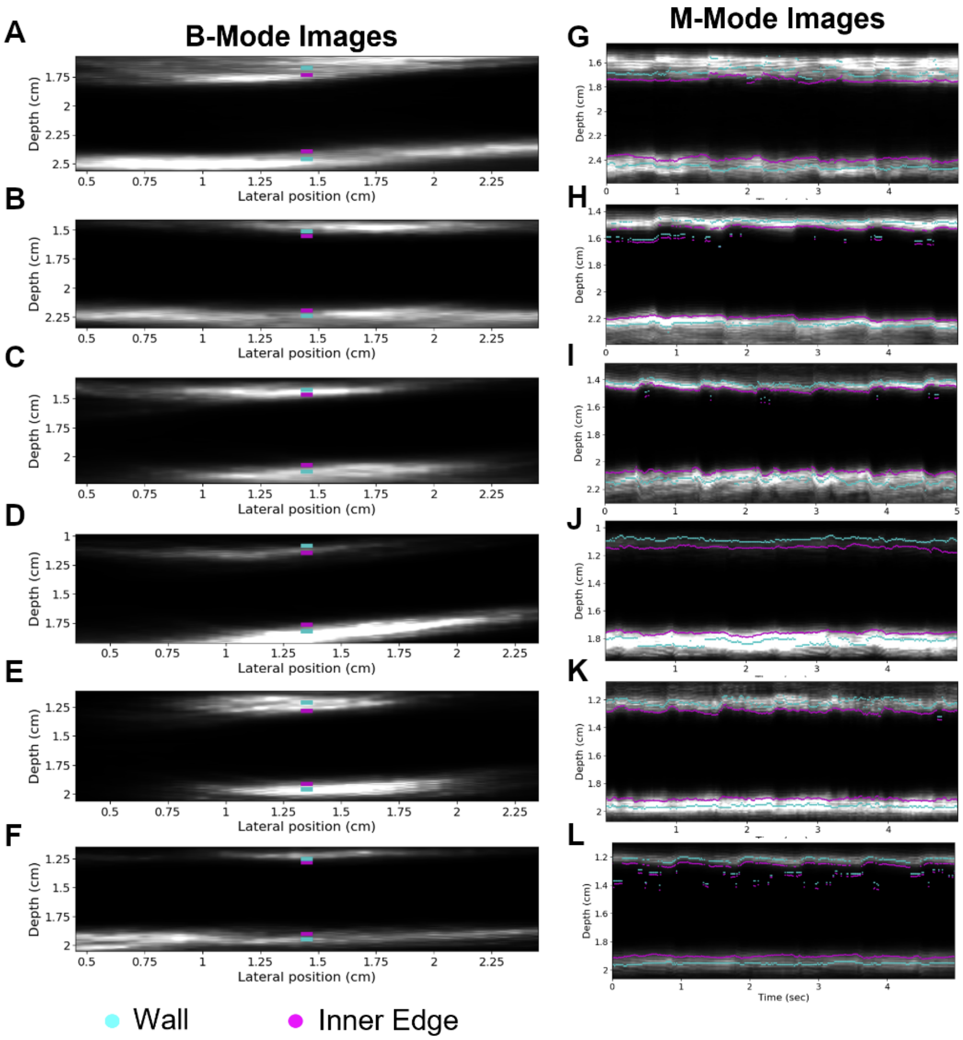

Figure S3: **Identification and tracking of vessel wall boundaries in time. A-F** on the left each depict an ultrasound B-mode image (five second average intensity projection) of the carotid artery in a longitudinal orientation corresponding to Subjects A-F in the $N = 6$ study, respectively. Wall velocity was calculated at the pixel column, where the magenta and cyan markers are located. **G-L** are corresponding raster images of the same five seconds of B-mode data, where the center 20 columns from each B-mode image were averaged to generate a single column for each timepoint (similar to processing for M-mode display). Magenta and cyan lines depict raw traces of the inner artery edge and interior wall (before radius filtering), respectively.

| Method | Modality | As presented by... | Calibration source | Full waveform? | Uses machine learning | Physical model? | Additional drawbacks/remarks |
|---|---|---|---|---|---|---|---|
| Ultrasound-imaged Resonance | Ultrasound | This manuscript | **None** | Yes | No | Yes (**fully determined**) | |
| Radius tracking | | Wang (2018) | Brachial cuff | Yes | No | No | Core exponential equation is empirical |
| Pulse-wave velocity (QA method) | | Seo (2021), Beulen (2011), Vappou (2011) | Finger cuff (Seo (2021)), brachial cuff (others) | Yes | No | Yes | Physical model uses Moens-Korteweg/Bramwell-Hill |
| Blood velocity wave analysis | | Jana (2020) | None / Brachial cuff (for training) | No | Yes (determines Windkessel model params.) | No* | *Windkessel provides underlying model, but feature extraction relies on linear regression. Use of machine learning results may result in misreporting during regimes outside of training set (such as abnormal/rare physiologies and cardiovascular shock) |
| Force-measured distension | | Zakrzewski (2018) | None | No | Yes | Yes | Requires steady applantation pressure, operator training required |
| Volume clamp | Finger cuff | Imholz (1998) | Brachial cuff (optional but recommended) | Yes | No | Yes | Difficult to measure BP on patients with low perfusion in extremities, periodic Physiocal self-calibration results in lower data availability |
| Volume control technique | | Fortin (2021) | Brachial cuff | Yes | No | No | Difficult to measure BP on patients with low perfusion in extremities |
| Pulse wave analysis | Capcitance measurement | Quan (2021) | Brachial cuff | No | Yes | No | |
| Pulse transit time analysis | Radar | Ibrahim (2019) | Brachial cuff | No | No | No – but BP from best fit of linear PTT model | |
| | PPG, ECG, video, etc. (review paper) | Mukkamala (2015) | Brachial cuff | No | No | Yes* | *Linear regression is used extensively to fit physics models) |
| Pulse wave analysis | Optical | Sola (2021) | Brachial cuff | No | No | No | |
| Amplitude correlation | PPG | Shaltis (2005) | Finger cuff | No | No | No | |
| Pulse wave analysis | Tonometer | Takazawa (2007) | Brachial cuff | No | Yes | No | Sensitive to noise and movement artifacts |
| Tonometry | Radar | Liao (2021) | Brachial cuff | Yes | No | No | |
| Pulse wave analysis | Bioimpedance | Ibrahim (2019) | Finger cuff | No | Yes | No | |
| Ballistocardiography (Pulse wave analysis) | Force plate | Kim (2018) | Finger cuff | No | No | No | Requires patient to stand/sit on force plate, not appropriate for ambulatory use |

Table S1: Comparison of noninvasive blood pressure measurement methods.

| Subject | Median Blood Pressure CV | Median Young's Modulus CV |
|---|---|---|
| A | 1.0e-4 | 1.1e-3 |
| B | 5.5e-5 | 5.0e-4 |
| C | 4.3e-5 | 4.9e-4 |
| D | 1.4e-5 | 1.4e-4 |
| E | 2.2e-5 | 2.4e-4 |
| F | 2.8e-5 | 3.9e-4 |

Table S2: Summary of our empirical investigation into the convergence of pressure and stiffness estimates from a range of initial conditions.

| Characteristic | | Data (n=6) |
|---|---|---|
| Age (yrs) | mean (range) | 33 (26-37) |
| Height (m) | mean (range) | 1.76 (1.60-1.85) |
| BMI (kg/m$^2$) | mean (range) | 24.03 (20.23-34.28) |
| Gender | Males - n (%) | 5 (83.3%) |
| | Females - n (%) | 1 (16.7%) |
| Race | Asian - n (%) | 1 (16.7%) |
| | White - n (%) | 4 (66.7%) |
| | Not Reported - n (%) | 1 (16.7%) |

Table S3: Demographics of six subject study.
